# Operationalizing Eight-Dimensional Patient-Safety Risk Scoring at Scale: A Multi-Model Large Language Model Reliability Study

**DOI:** 10.64898/2026.05.29.26354437

**Authors:** Hsuan-Ming Lin, JrJung Lyu, I-Lin Wang

## Abstract

**Background:** Hospital incident risk scoring has long relied on two-or three-dimensional frameworks (Severity Assessment Codes or Risk Priority Numbers), even though root cause analysis standards recognize that clinical risk is multi-factorial. The obstacle has been mainly cognitive: human reviewers cannot reliably score many dimensions across high incident volumes, so richer assessment has not been operationalized at scale.

**Objective:** To extend the traditional three-dimensional FMEA to an eight-dimensional patient-safety risk feature framework, to establish a multi-model large language model (LLM) extraction pipeline that scores these dimensions automatically, and to demonstrate a variance-aware integer optimization (mean-variance integer programming, MV-IP) that provides a reproducible tie-breaking rule for incident prioritization under extraction uncertainty, rather than improved risk coverage.

**Methods:** An 8-dimensional framework covering harm severity, potential harm, frequency, detectability, systemic impact, vulnerable populations, regulatory relevance, and economic impact was applied to 213 synthetic and 196 real curated incident narratives. Three independent LLMs (GPT-5.4, Gemini 3.1 Pro, Grok-4.1 Fast) from different provider families extracted structured risk scores. Inter-model consistency was assessed via ICC(A,1). Among coverage-equivalent selections, MV-IP minimized inter-model variance to give a reproducible prioritization rule. An English-language sensitivity analysis was conducted on 31 AHRQ PSNet WebM&M cases.

**Results:** On real cases, seven of eight dimensions reached Fair or better inter-model reliability (ICC(A,1) 0.53 to 0.83); D5 (Systemic Impact) was the exception at Poor reliability (0.275), driven by little between-case variation rather than by wide model disagreement. Reliability was not uniform: two dimensions were Excellent (D1 actual harm 0.834, D8 economic impact 0.782), two Good, and three only Fair, so some dimensions are more readily extractable than others. The same anchors gave broadly similar results on English-language narratives. When deterministic top-***Κ*** selection returned several equal-coverage solutions (11 on real cases, total inter-model variance 0.205 to 1.274), MV-IP selected the minimum-disagreement set, replacing ad hoc tie-breaking with an explicit rule without improving coverage. Bootstrap resampling found 74% to 90% of per-case variance estimates stable despite the three-model panel.

**Conclusions:** The eight-dimensional framework operationalizes patient-safety risk features that quality teams have considered only implicitly, and three independent LLM families produced reproducible scores on most dimensions of curated narratives. Inter-model agreement, however, measures reproducibility rather than clinical correctness, and high agreement does not by itself establish that a score is right; the dimensions that are reliably extractable today (notably D6 and D8) differ from those that are not yet (D5, and to a lesser degree D4 and D7), which has direct implications for incident-reporting form design. MV-IP contributes a reproducible, variance-aware tie-breaking rule rather than improved coverage. Validation against expert-prioritized RCA lists and deployment on raw institutional incident reports remain the next steps toward clinical use.

## 1 Introduction

### 1.1 Background

Current patient-safety risk tools can classify harm severity and probability, but cannot reliably distinguish incidents that share the same classification cell or numeric score while differing in systemic, regulatory, vulnerable-population, and economic risk. The **Severity Assessment Code (SAC)** matrix evaluates incidents along two dimensions (severity and probability) using a 4 × 4 grid developed by the U.S. Veterans Affairs National Center for Patient Safety [1]: widely adopted, yet unable to differentiate incidents within the same cell. The **Risk Priority Number (RPN)**, derived from Failure Mode and Effects Analysis (FMEA), adds a third dimension (Detection), but suffers from well-documented mathematical flaws: identical scores from fundamentally different risk profiles (e.g., 2 × 3 × 4 = 3 × 2 × 4 = 24), implicit equal weighting, and ordinal-scale multiplication that violates measurement theory [2, 3]. Two decades of alternatives, including fuzzy logic, MCDM, and AI-based approaches, have been proposed, yet most remain limited to the original three dimensions [4]. The Health-care FMEA (HFMEA) further reduced dimensionality by removing Detection entirely, acknowledging that “healthcare professionals cannot reliably assess detectability” [1]. These tool limitations matter at scale. Patient-safety incident reporting systems generate thousands of unstructured narratives annually across healthcare institutions worldwide, yet reporting systems improve safety only when incident data are translated into investigation, feedback, and system-level action [5]. National reporting data illustrate the scale of this workload [6]; because formal root cause analysis (RCA) is resource-intensive, institutions must stratify which events warrant full investigation [7, 8]. With current 2–3 dimensional tools unable to distinguish incidents that differ on systemic, regulatory, or economic axes, high-volume triage remains both cognitively demanding and methodologically constrained.

### 1.2 The Dimensionality Gap

The restriction to 2–3 dimensions does not reflect the true structure of clinical risk. Patient-safety professionals already *implicitly* consider systemic factors [9, 10], vulnerable populations, regulatory implications [11], and economic impact during RCA meetings. Both the Institute for Healthcare Improvement and the Canadian Patient Safety Institute have questioned the term “root cause analysis” itself, noting that “the term implies that there is one root cause, which is counter to the fact that health-care is complex and that there are generally many contributing factors” [7, 8]. The Canadian Incident Analysis Framework recommends assessing contributing factors across patient assessment, training, equipment, communication, policies, safety barriers, and patient characteristics [8]. The 2–3 dimensional FMEA frameworks therefore underspecify the multi-factorial structure that the RCA community itself recognizes. Why has this dimensionality gap persisted? A primary reason is the cognitive bottleneck: even with only three dimensions, inter-rater reliability in FMEA scoring is often poor [11]. Asking human reviewers to reliably score 8 dimensions across 50–70 monthly incidents is operationally infeasible. Large language models (LLMs) offer a potential solution to this cognitive constraint.

### 1.3 LLM-Assisted Multi-Dimensional Extraction

Recent advances in LLM-based clinical natural language processing have demonstrated the feasibility of zero-shot and few-shot information extraction from unstructured medical text [12, 13], while structured prompt engineering has been shown to improve output consistency [14, 15]. El Hassani et al. [16] demonstrated LLM-FMEA integration for industrial applications, but their approach relied on a single model family for both generation and evaluation, creating circular validation risk from self-preference bias [17], retained the three-dimensional RPN, and provided no external validation.

This study proposes to overcome both the dimensionality gap and the cognitive bottleneck simultaneously: extending the traditional 3-dimensional FMEA to an 8-dimensional risk feature framework, and using multiple independent LLMs from different provider families to extract these features at scale from unstructured incident narratives.

### 1.4 From Extraction to Prioritization Under Uncertainty

Reliable extraction alone does not solve the operational question: *which incidents should receive RCA when capacity is constrained?* Traditional MCDM methods such as TOPSIS [18] produce rankings but do not address resource constraints. A deterministic top-*K* selection treats extracted scores as exact, ignoring inherent LLM assessment uncertainty.

A mean-variance integer programming (MV-IP) formulation addresses this gap. Each incident is characterized by its median risk score (*µ_i_*) and inter-model variance (*σ*^2^). Drawing on the Markowitz portfolio optimization framework [19], MV-IP maximizes risk coverage subject to an uncertainty budget, providing a reproducible and auditable rule for incident prioritization. When multiple incidents share equivalent coverage, MV-IP selects the set with the lowest inter-model disagreement, replacing ad hoc tie-breaking with an explicit rule. Low disagreement is used here only to choose among equal-coverage sets; it is not a judgment that incidents the models disagree about are less important. On the contrary, high-disagreement incidents are routed to a separate adjudication queue for expert review (Section 4.9), so that model uncertainty triggers escalation rather than removal.

### 1.5 Study Objectives

This study aims to:

1. Extend the traditional 3-dimensional FMEA to an 8-dimensional patient-safety risk feature framework that formalizes dimensions previously considered only implicitly in clinical practice;
2. Establish a multi-model LLM extraction pipeline using independent model families and assess inter-model consistency on both synthetic and real incident cases;
3. Demonstrate a variance-aware integer optimization (MV-IP) that treats inter-model disagreement as a measure of extraction uncertainty and provides a reproducible rule for choosing among coverage-equivalent prioritizations under constrained RCA capacity; the aim is auditable tie-breaking, not improved risk coverage.

## 2 Methods

### 2.1 Study Design

This study employed a two-dataset design: 300 synthetic incident cases were generated, of which 255 were successfully loaded and 213 yielded complete extraction (Dataset A, see Case Inclusion Flow below), serving as a *development set* for method development, prompt optimization, and internal consistency assessment (prompt tuning used only SYN-P01 to P25; the primary synthetic reliability endpoint used final v1.2 extractions on the complete set, and all TPR results were external to prompt development); 198 publicly available real incident cases from a national patient-safety reporting system (Dataset B, NO.1 to 198, 2005 to 2025, 196 with complete extraction) served as the *external test set* assessing generalizability to curated authentic clinical narratives. Neither dataset required institutional review board approval: Dataset A consists of LLM-generated cases, and Dataset B comprises publicly released educational materials.

Inter-model consistency (ICC) validates extraction *reliability* (i.e., whether different models produce concordant scores), not clinical *validity* (i.e., whether scores are correct). Clinical validity against expert consensus is planned as a separate Phase 1.5 study (Section 4.10).

#### 2.1.1 Case Inclusion Flow

**Dataset A (synthetic):** 300 cases generated → 255 loaded (45 excluded: YAML parsing failures from malformed LLM output) → 213 with complete extraction across all 3 evaluators and all 8 dimensions (42 excluded: partial parse failures where ≥1 evaluator failed to return valid scores). Attrition details are provided in Supplementary Table S1. A goodness-of-fit *χ*^2^ test found no evidence that loading failures were systematically associated with event category (255 loaded vs. 300 target proportions; *χ*^2^=0.73, df=6, *p*=0.994). Category-specific extraction failure counts (255→213) could not be formally assessed because extraction outputs lack category labels; failure modes observed in parser logs were primarily malformed YAML or missing dimensions.

**Dataset B (real TPR):** 198 publicly available cases (NO.1–198, 2005–2025) → 196 with complete extraction across all 3 evaluators and all 8 dimensions (2 excluded due to ≥1 evaluator returning incomplete scores). Category distribution of the 198-case pool: medication 68 (34%), other 61 (31%), tube/line 23 (12%), surgery 22 (11%), fall 11 (6%), laboratory 10 (5%), transfusion 3 (2%); the 2 excluded cases do not change any category proportion by more than 0.5%.

#### 2.1.2 Sample Size

For ICC estimation with *k*=3 raters, a minimum of *n*=35 subjects is required to achieve 80% power for detecting ICC ≥ 0.75 against a null of ICC = 0.40 at *α*=0.05 (two-sided), based on the approximation by Bujang and Baharum [20]. Both Dataset A (*n*=213) and Dataset B (*n*=196) substantially exceed this minimum, providing adequate statistical power for per-dimension ICC estimation with narrow confidence intervals.

### 2.2 Eight-Dimensional Risk Feature Framework

This study extends the traditional FMEA three-dimensional framework (S×O×D) to eight dimensions (Table 2), grounding the extension in Donabedian’s Structure, Process, and Outcome quality assessment framework [21]. Dimensions D1 to D3 derive from established FMEA factors. D4 (Detectability) was restored via LLM extraction, previously removed by HFMEA due to human assessment limitations [1]. D5 to D8 formalize implicit considerations supported by Reason’s Swiss Cheese Model [9], WHO ICPS contributing factors and key concepts [22, 23], and Rezaei et al.’s revised RPN sub-dimensions [11]. Each dimension uses a 1 to 5 Likert scale with operationally defined anchors.

The eight dimensions were further mapped to the Triggering Questions for Root Cause Analysis recommended in the Canadian Incident Analysis Framework [8] and aligned with national patient-safety reporting system learning objectives [24] (Table 1), grounding the framework in established patient-safety practice rather than ad hoc theoretical choice. The Incident Decision Tree’s substitution test (“would another person make the same error?”) is operationalized in the D5 anchor for systemic-level scoring (D5≥4 when substitution suggests system-design failure rather than individual error).

**Table 1.** Eight-dimensional framework cross-walk to RCA triggering questions.

| RCA Triggering Question | Corresponding 8-dim dimension(s) |
| --- | --- |
| Q1: Patient assessment | D1 actual harm, D2 potential harm |
| Q2: Staff training/competency | D5 systemic impact (knowledge/skills layer) |
| Q3: Equipment | D5 (system layer) |
| Q4: Information sufficiency | D4 detectability, D5 |
| Q5: Communication | D5 (cross-team coordination) |
| Q6: Policies/procedures | D5, D7 regulatory relevance |
| Q7: Safety mechanisms (barriers) | **D4 detectability** (forcing functions) |
| Q8: Patient characteristics | **D6 vulnerable population** |

Triggering Questions based on the Canadian Incident Analysis Framework [8] and aligned with national reporting system learning objectives [24]. The framework operationalizes these assessment dimensions into machine-extractable Likert scales.

**Table 2.** Eight-dimensional risk feature framework.

| # | Dimension | Operational Definition | Source |
| --- | --- | --- | --- |
| D1 | Actual Harm Severity | Actual patient harm from the event | FMEA-S / SAC |
| D2 | Potential Harm Severity | Maximum harm if unintercepted | SAC close call |
| D3 | Base Frequency | Historical occurrence rate | FMEA-O |
| D4 | Detectability | Difficulty of detection (5=hardest)* | FMEA-D (restored) |
| D5 | Systemic Impact | Individual error vs. system flaw | Reason (1990) |
| D6 | Vulnerable Population | High-risk patient involvement | CMS Guidance |
| D7 | Regulatory Relevance | Mandatory reporting / legal risk | Rezaei et al. (2018) |
| D8 | Economic Impact | Additional treatment magnitude | Rezaei et al. (2018) |

\*All dimensions are scored so that 5 = highest priority / highest risk. D4 uses reverse coding (5 = hardest to detect = highest risk), consistent with traditional FMEA convention where low detectability indicates higher priority.

### 2.3 Multi-Model LLM Extraction

#### 2.3.1 Model Separation Design

To avoid circular validation bias and self-preference bias documented in LLM evaluation literature [16, 17], we employed strict model separation: the Generator (Claude Opus/Sonnet, Anthropic) produced synthetic cases and designed extraction prompts, while three independent Evaluators from different model families extracted risk features (Table 3). This cross-model consistency approach treats independent LLM families as separate coders, providing a reliability signal analogous to inter-coder agreement in computational linguistics [25].

**Table 3.** Model roles and separation design.

| Role | Model | Provider | Purpose |
| --- | --- | --- | --- |
| Generator | Claude Opus 4 / Sonnet 4 | Anthropic | Case generation + prompt design |
| Evaluator 1 | GPT-5.4 | OpenAI | Independent 8-dim extraction |
| Evaluator 2 | Gemini 3.1 Pro | Google | Independent 8-dim extraction |
| Evaluator 3 | Grok-4.1 Fast | xAI | Independent 8-dim extraction |

All extraction runs used temperature=0.1, structured YAML output, and single-pass extraction (no retry after a valid API response). Because provider routes and output-token settings differed by model and dataset, run provenance is summarized in Table 4 rather than treated as a single uniform configuration. Parse failures were excluded from the complete-case reliability analysis and are reported as a pipeline performance endpoint. API access dates: April 2026. Commercial LLM APIs do not guarantee version-pinning; providers may update model weights without changing identifiers. Exact reproducibility therefore cannot be guaranteed. The complete extraction prompt (v1.2), system prompt, parse rules, model/run provenance appendix, and analysis code are available from the corresponding author on reasonable request.

**Table 4.**
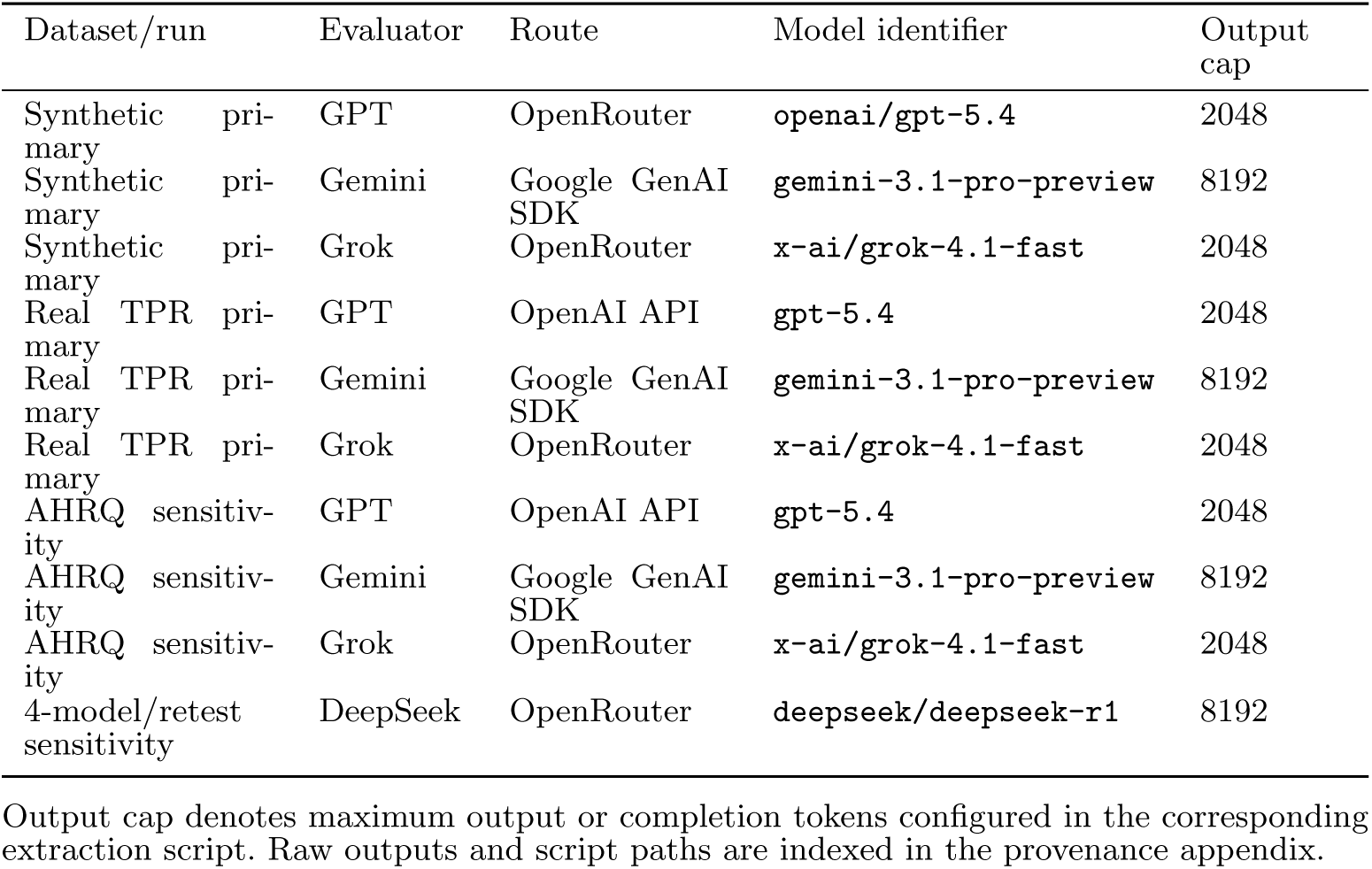
Model route and run-configuration provenance for primary and sensitivity extraction runs.

Final scores were determined by median aggregation across the three evaluators.

#### 2.3.2 Prompt–Model Development Trajectory

A prompt–model development trajectory was adopted, examining each dimension’s inter-model consistency across successive prompt and evaluator configurations (Table 5). **Important caveat:** versions v1.0, v1.1, and v1.2 differed not only in prompt content but also in model version and sample size; observed changes should therefore be interpreted as development trajectory observations rather than causal attributions to prompt changes alone. Prompt development used only the 25-case pilot pool (SYN-P01 through SYN-P25). The final v1.2 synthetic reliability run attempted all 255 successfully loaded synthetic cases, including the pilot cases re-extracted under the final prompt; thus the primary synthetic ICC endpoint is a final-prompt complete-case estimate, not an untouched holdout estimate. The 188 non-pilot complete cases were never used to select or modify the prompt, and all real-case TPR results were external to prompt development. This design limits, but does not eliminate, prompt-tuning leakage risk. All real-case TPR results (*n*=196) are fully external to prompt development and serve as the primary reliability benchmark; the synthetic ICC should be interpreted as a development-set estimate. A non-pilot-only synthetic sensitivity analysis is a potential future extension but is not required for the primary conclusions, which rest on external real-case data.

**Table 5.** Prompt–model development trajectory. Note: versions differ in prompt content, model version, and sample size; improvements are trajectory observations, not causal attributions.

| Ver | Pool | Valid $n$ | Issue | Prompt addition | Observed change |
| --- | --- | --- | --- | --- | --- |
| v1.0 | 25 | 11 | D3 ICC=0.567 | Frequency ref. table (20 subtypes) | D3→0.831 |
| v1.1 | 75 | 20 | D5 Gemini bias 38% | Systemic-level calib. guide + pitfalls | Bias→4% |
| v1.2 | 255 | 213 | Production | – | All dims $\geq 72\%$ W-1 |

Pool = cases attempted; Valid $n$ = cases with complete extraction across all evaluators used for ICC calculation (Table 6).

The development trajectory suggested that two types of prompt augmentation may be useful: (1) *structured reference tables* providing domain-specific factual knowledge (for inference-dependent dimensions like frequency), and (2) *calibration examples* with decision rules and common pitfalls (for judgment-dependent dimensions like systemic impact). Because prompt content, evaluator model, and sample size changed simultaneously, these patterns should not be interpreted as isolated prompt effects.

To document the per-dimension trajectory across prompt versions, Table 6 reports ICC values for all eight dimensions under v1.0, v1.1, and the v1.2 production run. Two patterns are notable: (a) D3 (base frequency) changed from 0.567 to 0.896 after addition of the structured reference table in v1.1; (b) D5 (systemic impact) showed an apparent ICC reduction after v1.2 calibration additions (0.701 → 0.539), with the Gemini D5=5 over-rating rate dropping from 38% to 4% and scores migrating toward the modal value of 3. These patterns are descriptively useful, but inter-version effects should be interpreted as joint prompt × model effects because v1.0/v1.1 used lighter evaluator models (GPT-4.1, Gemini 2.5 Flash, Grok-3-mini) while v1.2 production used flagship models; pure prompt effect is not isolated.

**Table 6.**
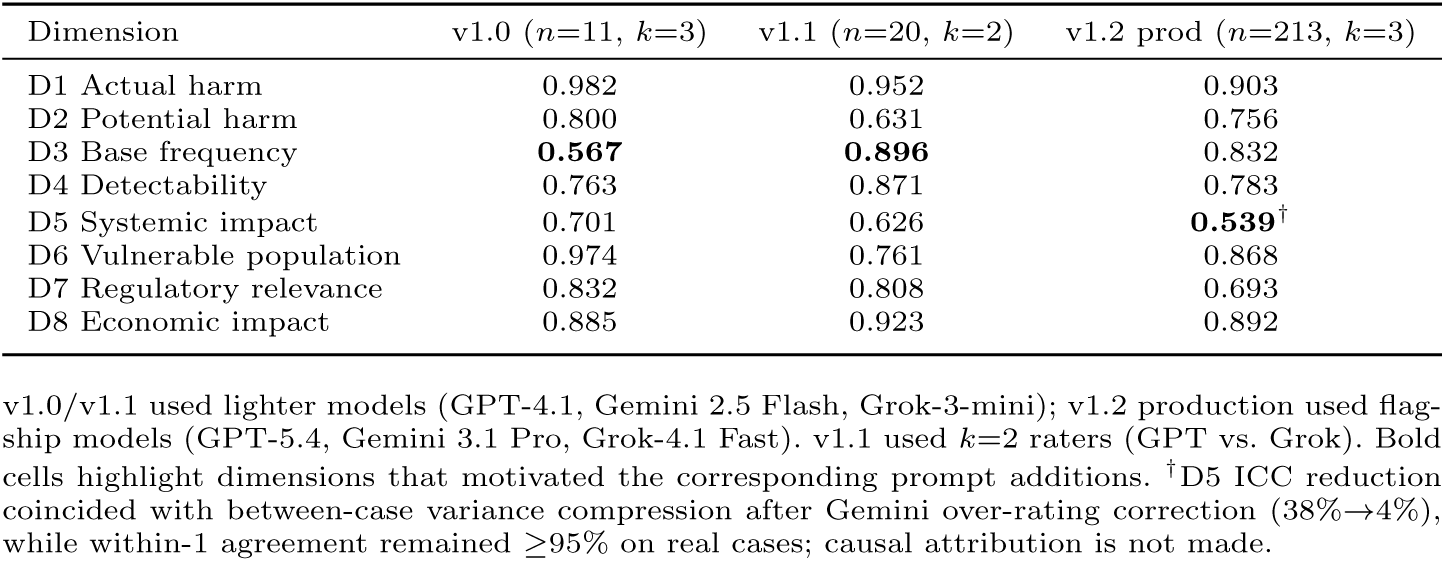
Per-dimension ICC(A,1) across prompt versions (synthetic data).

#### 2.3.3 Inter-Model Consistency Assessment

Consistency was assessed using dual metrics following Koo & Li [26] and GRRAS guidelines [27]: ICC(A,1), two-way random-effects, absolute agreement, single measures [28], for dimensions with sufficient between-case variance, and within-1 percentage agreement for restricted-range dimensions. The two-way random model was selected because evaluators are treated as random effects (not fixed to these specific three models), and absolute agreement, rather than consistency, is required because scores must be interchangeable across evaluators [29]. This dual-reporting approach addresses the known paradox where ICC underestimates reliability in homogeneous samples [30]. ICC benchmarks follow Cicchetti’s guidelines: *<*0.40 poor, 0.40 to 0.59 fair, 0.60 to 0.74 good, ≥0.75 excellent [31]. Given 16 primary ICC tests (8 dimensions × 2 datasets), the Bonferroni-corrected significance threshold is *α*=0.05/16=0.003. All 16 primary ICCs were significant at the corrected threshold; exact *p* values are reported in Supplementary Tables. The smaller AHRQ sensitivity sample (*n*=31) yields wider confidence intervals and is not included in the multiplicity correction. Following psychometric convention, we emphasize 95% confidence intervals rather than *p* values as the primary uncertainty quantification throughout [26].

#### 2.3.4 Supplementary Agreement Metrics and Reliability-Paradox Diagnostics

In addition to ICC(A,1), the following were computed: (a) within-1 percentage agreement averaged over evaluator pairs to provide a clinically interpretable reliability measure when ICC is suppressed by restricted score range, and (b) the modal-score concentration per dimension as a diagnostic for the kappa-or ICC-paradox [30], a phenomenon in which chance-corrected reliability statistics are deflated by homogeneous marginal distributions despite high underlying agreement. Cohen’s *κ* is not reported as the primary reliability metric because *κ* assumes nominal scales and is known to be unstable under restricted-range ordinal distributions for the same paradox reasons [26, 30].

#### 2.3.5 Sensitivity Analyses

*Four-model robustness check.* To examine whether our reliability findings depend on the specific choice of three evaluators, we additionally extracted scores using DeepSeek R1 (a Chinese open-source reasoning model from a fourth provider family) and computed ICC with *k*=4 raters on cases with complete 4-model extraction (synthetic *n*=199; real *n*=194 after collapsing lab/laboratory category-label variants). This is reported as supplementary sensitivity rather than primary analysis to preserve methodological consistency with the pre-specified 3-model design.

*Test-retest reliability.* To assess within-model stochastic noise from temperature=0.1, a random sample of 30 real cases (seed=42, no stratification) was re-extracted by each of the four evaluators in a second independent run. Within-model ICC(A,1) between run 1 and run 2 was computed per dimension. This is exploratory; the small sample and lack of category stratification preclude population-level conclusions.

*Prompt* × *model joint effect.* The prompt iteration ablation (Table 6) compares v1.0/v1.1 against v1.2 production. Because v1.0/v1.1 used lighter evaluator models (GPT-4.1, Gemini 2.5 Flash, Grok-3-mini) on a smaller pilot set while v1.2 production used flagship models on the full set, the inter-version trajectory reflects a joint prompt × model effect rather than pure prompt effect. Re-running v1.0/v1.1 prompts on flagship models was not feasible due to API cost on the full sample. The Model Capacity Ablation Study (9 2.3.6) addresses the model-effect contribution directly with a controlled comparison on the same v1.2 prompt.

*External English-language narrative sensitivity analysis.* To examine whether the framework’s reliability patterns were limited to Chinese-language TPR narratives, we extracted 31 publicly-available English-language patient safety cases from the AHRQ Patient Safety Network (PSNet) WebM&M repository [32]. Cases were obtained directly via raw HTTP from PSNet (no LLM pre-processing) and span medication, fall, surgery, communication, diagnostic, and other categories. The same v1.2 Chinese-language extraction prompt was applied unchanged to each English narrative; we did not translate or rewrite the prompt, in order to explore whether the same scoring anchors could be applied to an English-language public case corpus rather than testing a translated prompt’s equivalence. Per-dimension ICC(A,1) was computed across the same three flagship evaluators (GPT-5.4, Gemini 3.1 Pro, Grok-4.1 Fast) and compared dimension-by-dimension to the TPR Chinese real-case ICC. This analysis is interpreted as a narrative-sensitivity check because language, case genre, and narrative richness are confounded.

#### 2.3.6 Model Capacity Ablation Study

To assess whether inter-model consistency depends on model capacity or prompt design, we conducted an ablation study comparing flagship models (GPT-5.4, Gemini 3.1 Pro, Grok-4.1 Fast) against lighter models (GPT-4.1, Gemini 2.5 Flash, Grok-3-mini) on the same 55 real TPR cases using the same v1.2 prompt.

### 2.4 Mean-Variance Integer Optimization (Module B)

#### 2.4.1 Problem Formulation

Incident prioritization is formulated as a mean-variance integer optimization problem, using Markowitz portfolio selection [19] as a mathematical analogy. The clinical interpretation is narrower: preserve risk coverage first, then minimize model-disagreement variance among coverage-equivalent selections. Each incident *i* is characterized by:

- *µ_i_*: median risk aggregate across three evaluators (return analog)
- *σ*_i_^2^: inter-model variance of risk aggregate (risk analog)

The optimization problem is:

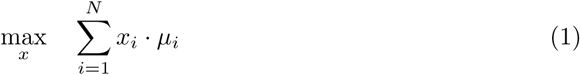

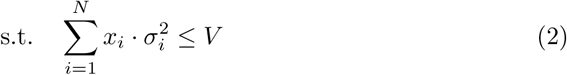

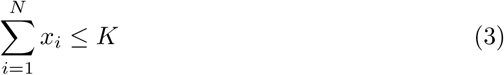

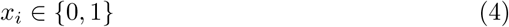

where *V* is the uncertainty budget and *K* is the monthly RCA capacity (set to 10 based on institutional practice). A category-coverage constraint of the form Σ*_i_*_∈_*_Cj_ x_i_* ≥ 1 for each event category *C_j_* can be added as an optional extension; we report results without this constraint to keep the optimization core minimal, and assess category diversity post-hoc as one of the comparison metrics in Table 13.

The risk aggregate *µ_i_*was computed as the equal-weight average of 8 dimensions:

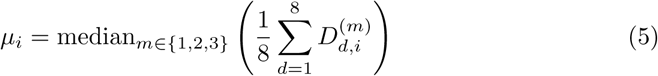

and the uncertainty parameter:

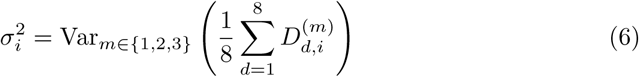

where *D_d,i_*^(*m*)^ denotes the score assigned by evaluator *m* to dimension *d* of incident *i*. The composite *µ_i_* is a pragmatic index for the prototype prioritization model, not a validated interval-scale utility measure. The primary reliability analyses are reported per dimension and do not depend on this equal-weight composite. Expert-derived utility weights and ordinal multi-criteria decision methods are planned for validation-phase work.

We note candidly that averaging eight ordinal Likert scores into the composite *µ_i_* shares the measurement-theoretic limitation we raise against RPN in Section 1.1, namely arithmetic on ordinal scales. We use *µ_i_* only as a screening index for the prototype selection step, not as a clinical priority score, and the per-dimension reliability results that constitute the study main findings do not depend on it. Replacing the equal-weight average with expert-derived weights and ordinal multi-criteria methods (for example the Best-Worst Method) is part of the planned validation phase. Because Datasets A and B are static curated corpora rather than a single institution monthly intake, the optimization is presented as a demonstration of the selection rule on a fixed pool; *K* equal to 10 should be read as an illustrative capacity, not as a literal monthly queue applied to these specific cases.

#### 2.4.2 Efficient Frontier

By sweeping *V* from tight (only high-confidence incidents) to loose (equivalent to deterministic knapsack), we trace an efficient frontier revealing the trade-off between risk coverage and extraction reliability. The recommended operating point was identified using the elbow method: the value of *V* where marginal coverage gains diminish sharply.

#### 2.4.3 Independence Assumption

An independent variance assumption is adopted (Σ*x_i_σ_i_*^2^ ≤ *V*), treating each incident’s extraction uncertainty as independent. While a full covariance structure (**x***^T^* **Σx**) could capture cross-incident correlations in model disagreement, the *N* × *N* covariance matrix is severely underdetermined with only *k* = 3 model observations. This limitation is discussed in Section 4.10.

#### 2.4.4 Implementation

All optimization was performed using scipy.optimize.milp in Python 3.13 with the HiGHS solver. The problem size (*N* ≈ 213, *K* = 10) is tractable for exact solution.

#### 2.4.5 Optimization Weight Sensitivity

To assess the impact of equal-weight assumptions on MV-IP selection, we recomputed *µ_i_* under four alternative weighting schemes: equal (baseline), harm-heavy (D1=D2=0.20), regulatory-heavy (D7=0.25), and safety-first (D1=D2=D5=D6=0.15). For each scheme, the MV-IP optimization was re-solved and top-10 selection overlap was compared across schemes.

#### 2.4.6 Baseline Comparison

The MV-IP model was compared against five alternative selection methods: (1) Top-*K* by *µ* (deterministic knapsack), (2) Top-*K* by Lower Confidence Bound (*µ* − 1.5*σ*), (3) Pareto-front selection (non-dominated on *µ* and −*σ*^2^), (4) stratified random selection within event categories (1000 simulations), and (5) SAC-proxy ranking (D1 × D3). Comparison metrics included risk coverage (Σ*µ*), total uncertainty (Σ*σ*^2^), high-harm recall, and category diversity.

### 2.5 Datasets

#### 2.5.1 Dataset A: Synthetic Incident Cases

A total of 300 synthetic incident cases were generated reflecting hospital scenarios across 7 WHO ICPS categories (medication 30%, falls 15%, tube/line 10%, surgery 10%, laboratory 10%, transfusion 5%, others 20%). Cases were generated by Claude Opus (75 cases, manual quality review) and Claude Sonnet (225 cases, batch API) using an iteratively optimized generation prompt (v1.1) based on analysis of 55 TPR public cases. Of 300 generated cases, 255 (85.0%) were successfully loaded into the extraction pipeline (45 cases dropped at the YAML loading stage due to malformed structure or missing required fields). Of these 255 loaded cases, 213 (83.5% of loaded cases; 71.0% of generated cases) yielded complete extraction across all three evaluators and all 8 dimensions; the remaining 42 cases had parse failures from at least one evaluator (malformed YAML output) and were excluded from all primary complete-case reliability analyses. Thus, the reported ICCs characterize successful complete extractions, while the complete-extraction rate itself should be interpreted as an additional pipeline reliability endpoint. A production implementation should incorporate schema validation, retry/repair logic, and prospective monitoring of parse failures rather than treating failed extractions as ignorable missingness.

Synthetic cases generated by one LLM family (Claude) may exhibit characteristics that make them more predictable for other LLMs to score. A formal comparison confirmed this: real narratives were 3.1× longer (3,060 vs. 990 characters, *p <*0.001), had × higher inter-model variance (0.041 vs. 0.016, *p <*0.001), and showed significantly different score distributions on D4 and D7 (*p <*0.001). However, core harm dimensions (D1, D2, D5, D6) showed no significant distributional differences (*p >*0.05), indicating that the synthetic cases’ risk profiles, though easier to extract, are broadly comparable to real incident severity patterns within the reliability scope. Dataset B therefore serves as the primary external reliability benchmark.

#### 2.5.2 Dataset B: TPR Real Incident Cases

A total of 198 publicly available **curated educational TPR cases** (NO.1 to 198, spanning 2005 to 2025) were obtained from a national patient-safety reporting system’s public educational corpus. These are real clinical events that have been edited and standardized for educational dissemination: they are not raw incident reports but curated learning materials. Because these cases are publicly released as educational resources without identifiable patient information, analysis did not require institutional review board approval. API access for extraction was performed via standard commercial endpoints (OpenAI, Google AI Studio, OpenRouter); no identifiable patient information was submitted, and vendor data-use policies should be re-verified before any deployment involving institutional data. Categories: medication (34%), other (31%), tube/line (12%), surgery (11%), falls (6%), laboratory (5%), and transfusion (2%). Cases are consecutively numbered and represent the complete publicly available corpus of TPR learning cases and alerts. The term “real” is used throughout to denote authentic clinical content (vs synthetic cases generated by Claude), with the caveat that generalization to raw, uncurated institutional incident reports requires Phase 2 validation.

## 3 Results

### 3.1 Inter-Model Consistency

#### 3.1.1 Pipeline Completion and Parse Failures

Pipeline completion is reported separately from complete-case ICC because parse failure is part of full-system reliability. For Dataset A, 300 synthetic cases were generated, 255 (85.0%) were successfully loaded after YAML parsing, and 213 (71.0% of generated; 83.5% of loaded) yielded complete valid scores across all three primary evaluators and all 8 dimensions. Per-model parse failure rates among the 255 loaded synthetic cases were 21/255 (8.2%) for GPT-5.4, 41/255 (16.1%) for Gemini 3.1 Pro, and 0/255 (0.0%) for Grok-4.1 Fast. For Dataset B, 196 of 198 real TPR cases (99.0%) yielded complete valid scores; both exclusions were caused by structurally invalid Grok-4.1 Fast outputs, while GPT-5.4 and Gemini 3.1 Pro returned complete scores. Therefore, all ICCs below should be interpreted as conditional on complete valid extraction rather than as full-pipeline success rates. Detailed attrition and failure descriptions are provided in Supplementary Table S1.

#### 3.1.2 Synthetic Cases (Dataset A)

Among 255 synthetic cases, 213 had complete extraction across all three evaluators and all 8 dimensions (42 excluded due to parse failures; see Methods). Per-dimension results are shown in Table 7.

**Table 7.** Inter-model consistency on synthetic development cases (*n*=213, 3 evaluators).

| # | Dimension | ICC(A,1) [95% CI] | $\kappa_w$ | AC2 | Within-1 |
| --- | --- | --- | --- | --- | --- |
| D1 | Actual Harm | 0.903 [0.88, 0.92] | 0.902 | 0.902 | 98% |
| D2 | Potential Harm | 0.756 [0.71, 0.80] | 0.756 | 0.755 | 94% |
| D3 | Frequency | 0.832 [0.80, 0.87] | 0.832 | 0.832 | 100% |
| D4 | Detectability | 0.783 [0.73, 0.83] | 0.783 | 0.781 | 88% |
| D5 | Systemic Impact | 0.539<sup>†</sup> [0.46, 0.61] | 0.551 | 0.534 | 97% |
| D6 | Vulnerable Pop. | 0.868 [0.84, 0.89] | 0.867 | 0.867 | 93% |
| D7 | Regulatory | 0.693 [0.63, 0.75] | 0.691 | 0.692 | 96% |
| D8 | Economic Impact | 0.892 [0.86, 0.92] | 0.891 | 0.892 | 99% |

ICC(A,1) = two-way random, absolute agreement, single measures [28]. $\kappa_w$ = mean pairwise quadratic weighted kappa. AC2 = Gwet’s AC2. The near-identity of ICC, $\kappa_w$ , and AC2 is consistent with the known close relationship among these metrics: for equally-spaced ordinal scales with symmetric rater designs, ICC(A,1) and quadratic weighted kappa converge closely [33]; AC2 yields similar values when marginal distributions are not extremely skewed. D5 shows the largest divergence among the three metrics, consistent with its restricted marginals. <sup>†</sup>D5 (exploratory): restricted range; 87.6% of all ratings = score 3; 88.1% exact 3-way agreement. Low ICC is consistent with compressed between-case variance; large-magnitude disagreement was uncommon, but discriminative reliability was poor [26, 29]. D5 is retained as theoretically motivated (Reason’s Swiss Cheese Model) but interpreted as exploratory given insufficient discriminative power from brief narratives.

#### 3.1.3 Real TPR Cases (Dataset B)

Among 198 real TPR cases (NO.1 to 198, spanning 2005 to 2025), 196 had complete extraction across all three evaluators. Per-dimension results are shown in Table 8.

**Table 8.** Inter-model consistency on real TPR cases (*n*=196, NO.1 to 198, 2005 to 2025).

| # | Dimension | ICC(A,1) [95% CI] | Interpretation |
| --- | --- | --- | --- |
| D1 | Actual Harm | 0.834 [0.78, 0.87] | Excellent |
| D2 | Potential Harm | 0.565 [0.49, 0.64] | Fair |
| D3 | Frequency | 0.699 [0.63, 0.76] | Good |
| D4 | Detectability | 0.529 [0.45, 0.61] | Fair |
| D5 | Systemic Impact | 0.275<sup>†</sup> [0.18, 0.37] | Poor |
| D6 | Vulnerable Pop. | 0.745 [0.69, 0.79] | Good |
| D7 | Regulatory | 0.593 [0.52, 0.66] | Fair |
| D8 | Economic Impact | 0.782 [0.72, 0.83] | Excellent |

<sup>†</sup>D5 (exploratory): restricted range (majority of cases score 3); retained as theoretically motivated but not reliably discriminative from brief narratives. Seven of eight dimensions achieved Fair or better reliability ( $\geq 0.40$ ); only D5 fell below Fair. D1 and D8 achieved Excellent ( $\geq 0.75$ ), showing high inter-model reliability for harm severity and economic impact extraction from real clinical narratives.

The per-dimension ICC variation between synthetic and real cases reflects the greater clinical ambiguity inherent in authentic narratives. Notably, D1 (Actual Harm) achieved Excellent reliability (0.834) on real cases, indicating high inter-model consistency for concrete harm outcomes regardless of narrative complexity.

Figure 1 provides a unified visualization of per-dimension ICC across the three evaluation datasets (synthetic, TPR Chinese real, and AHRQ English; the AHRQ English-case sensitivity analysis is detailed in Section 3.5.3). Two patterns are visu-ally evident: (a) most dimensions fall in the Fair-to-Excellent range across all three datasets, and (b) D5 (systemic impact) is consistently the lowest-ICC dimension on all three, consistent with restricted-range effects and limited textual discrimination across datasets.

**Fig. 1.**
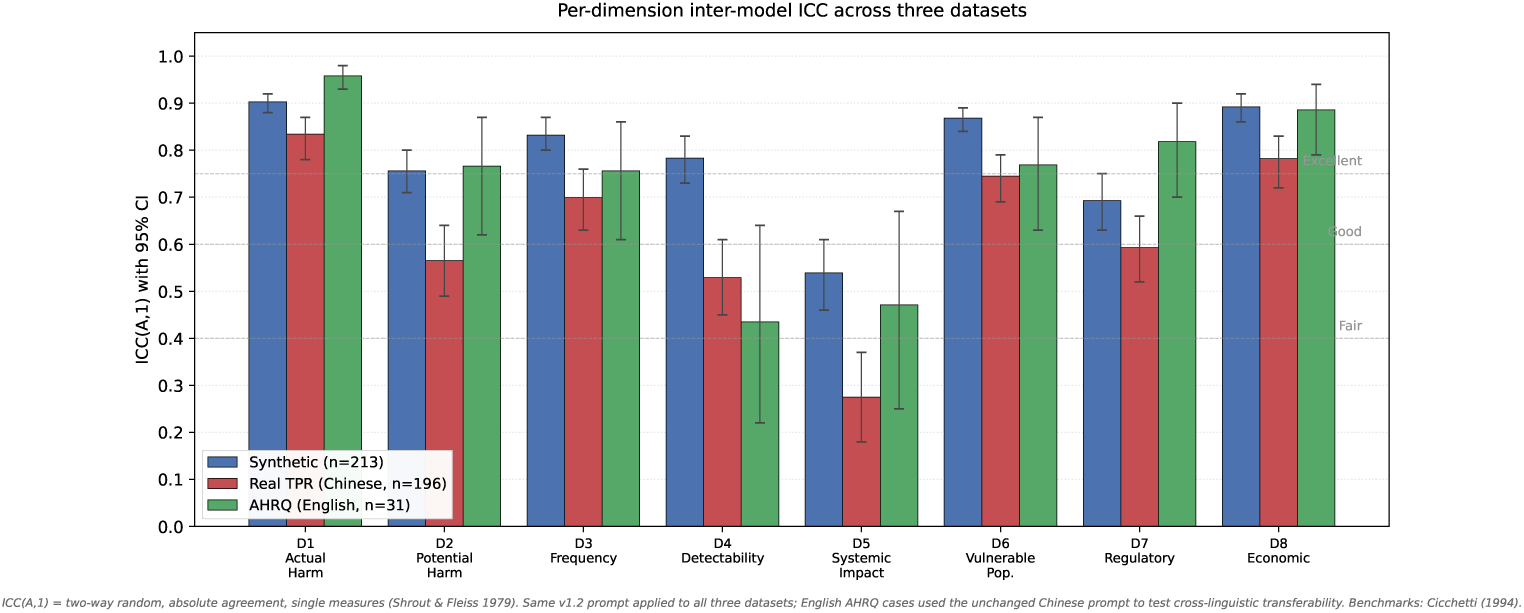
Per-dimension inter-model ICC(A,1) with 95% CI across three datasets. Cicchetti (1994) reliability benchmarks shown as horizontal reference lines. The same v1.2 prompt was applied to all three datasets; English AHRQ cases used the unchanged Chinese prompt. Seven of eight dimensions achieve Fair-to-Excellent reliability on the primary real TPR dataset; D5 (Systemic Impact) is the lowest-ICC dimension on all three datasets, consistent with restricted score range and limited textual discrimination (Section 4.5). D4 (Detectability) is the second-weakest on real data.

### 3.2 Prompt–Model Development Trajectory

Table 5 summarizes the prompt–model development trajectory, and Table 6 reports per-dimension ICC across v1.0, v1.1, and the v1.2 production run. The development trajectory showed two notable dimension-specific patterns:

1. **D3 (Frequency):** ICC increased from 0.567 (v1.0, *n*=11) to 0.896 (v1.1, *n*=20), coinciding with the addition of a structured frequency reference table with 20 event subtypes (Δ=+0.329). The v1.2 production result on *n*=213 was 0.832. Because prompt content, model version, and sample size changed simultaneously, the relative contribution of the reference table cannot be isolated from model-capacity effects.
2. **D5 (Systemic Impact):** After adding a systemic-level calibration guide, one evaluator’s outlier rate decreased from 38% to 4%. ICC numerically decreased (0.701 v1.0 → 0.539 v1.2), consistent with compressed between-case variance: when scores cluster tightly (74% to 90% at modal value 3 in v1.2), ICC declines mathematically regardless of agreement magnitude [26, 30]. Within-1 agreement on real cases remained ≥95%, suggesting that poor ICC reflects restricted score range and limited textual discrimination rather than frequent large-magnitude disagreement.

Prompt support needs differed by dimension. *Inference-dependent* dimensions (e.g. D3 frequency) benefited from factual reference tables, while *judgment-dependent* dimensions (e.g. D5 systemic impact) benefited from calibration examples and common-pitfall guidance. The v1.0/v1.1/v1.2 trajectory cannot fully isolate prompt effects from model effects, because v1.0/v1.1 used lighter evaluator models (GPT-4.1, Gemini 2.5 Flash, Grok-3-mini) while v1.2 production used the flagship trio; the ablation table reports the joint prompt × model effect, not pure prompt effect. The Model Capacity Ablation Study below addresses the model-effect contribution directly.

### 3.3 Model Capacity Ablation Study

The ablation study applied the same v1.2 prompt to 55 real TPR cases using both flagship (GPT-5.4, Gemini 3.1 Pro, Grok-4.1 Fast) and lighter (GPT-4.1, Gemini 2.5 Flash, Grok-3-mini) model panels. Complete-case denominators differed substantially between panels due to parse failures: flagship *n*=54 (98% complete) versus lighter *n*=32 (58% complete). Pooled ICC(A,1) across all 8 dimensions was 0.713 (flagship, *n*=54) versus 0.716 (lighter, *n*=32); per-dimension mean ICC was 0.580 versus 0.538, respectively (Supplementary Table S5). The flagship panel’s primary advantage was output format compliance rather than scoring consistency: parse success increased from 75% (lighter, averaged across 3 models) to 99% (flagship). This pattern is compatible with the possibility that structured prompt calibration contributed materially to inter-model consistency, while model capacity primarily improved output-format compliance in this 55-case comparison.

### 3.4 Variance-Aware Tie-Breaking Under Extraction Uncertainty

#### 3.4.1 Data Characteristics

Among 213 synthetic cases with complete extraction, the mean risk aggregate ranged from 1.88 to 4.25 (*µ*=2.86). Inter-model variance ranged from 0.0000 to 0.1354 (*σ*^2^=0.0157), with 26 cases (12%) showing perfect agreement (*σ*^2^=0).

Because *σ_i_*^2^ is estimated from only *k*=3 evaluators (df=2), we assessed estimation stability via nonparametric bootstrap (2000 resamples of evaluator panels with replacement per case, seed=42). On synthetic cases, 90.1% of per-case bootstrap 95%

CIs for *σ*^2^ had width *<*0.05, indicating that the variance estimates are stable for the large majority of cases despite the small *k*. On real TPR cases, 74.0% of CIs were narrow (*<*0.05); the remaining 26% showed wider CIs, identifying cases where MV-IP selection is sensitive to the specific evaluator panel. Full per-case bootstrap results are available in the project repository (bootstrap-sigma2-results.json).

#### 3.4.2 Efficient Frontier

Figure 2 shows the efficient frontier for *K*=10. Nine distinct operating points were identified. At the recommended operating point (*V* =0.058, identified via elbow method):

- Risk coverage decreased by only 1.0% (37.375 → 37.000)
- Total LLM uncertainty decreased by 82.0% (0.309 → 0.056)
- 4 of 10 selected incidents changed compared to the deterministic solution
- Selection overlap with pure knapsack: 60%

**Fig. 2.**
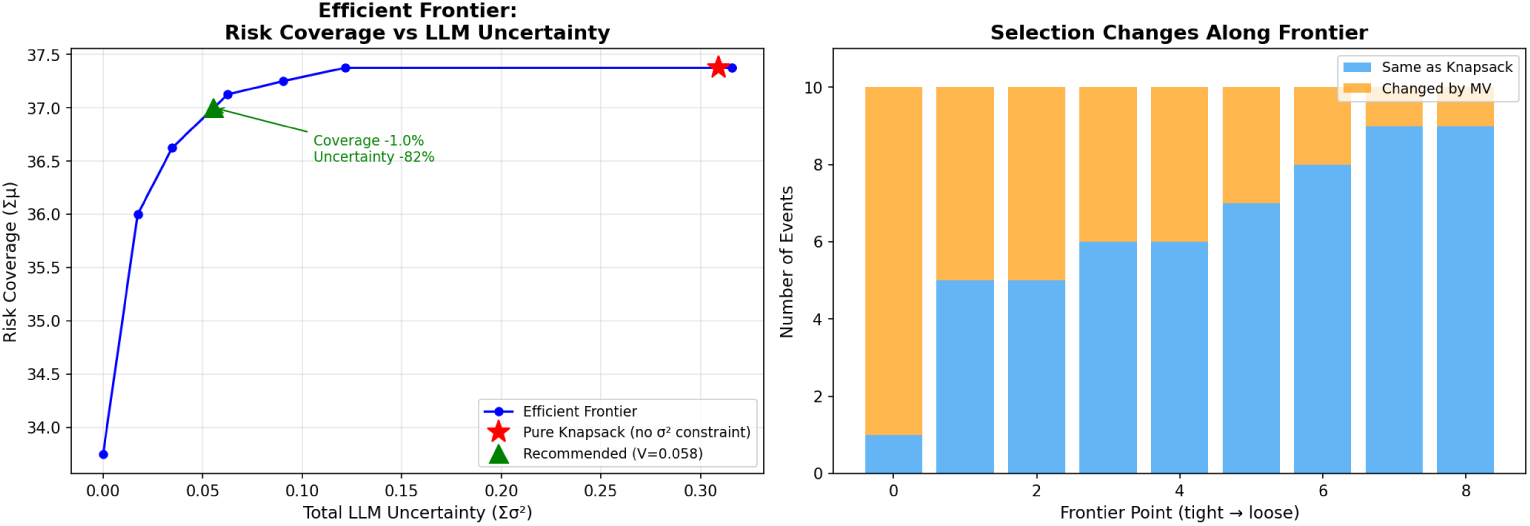
Efficient frontier: risk coverage versus LLM uncertainty. Red star = deterministic knapsack (no uncertainty constraint). Green triangle = recommended operating point (*V* =0.058). The efficient frontier reveals a clinically interpretable trade-off between coverage-maximizing (reactive) and reliability-prioritizing (conservative) strategies.

#### 3.4.3 Selection Changes

Figure 3 visualizes case selection in *µ*-*σ*^2^ space. The mean-variance model replaced 4 incidents with high *µ* but high *σ*^2^ (“high-scoring but contentious”) with incidents having slightly lower *µ* but near-zero *σ*^2^ (“slightly lower-scoring but reliable”).

**Fig. 3.**
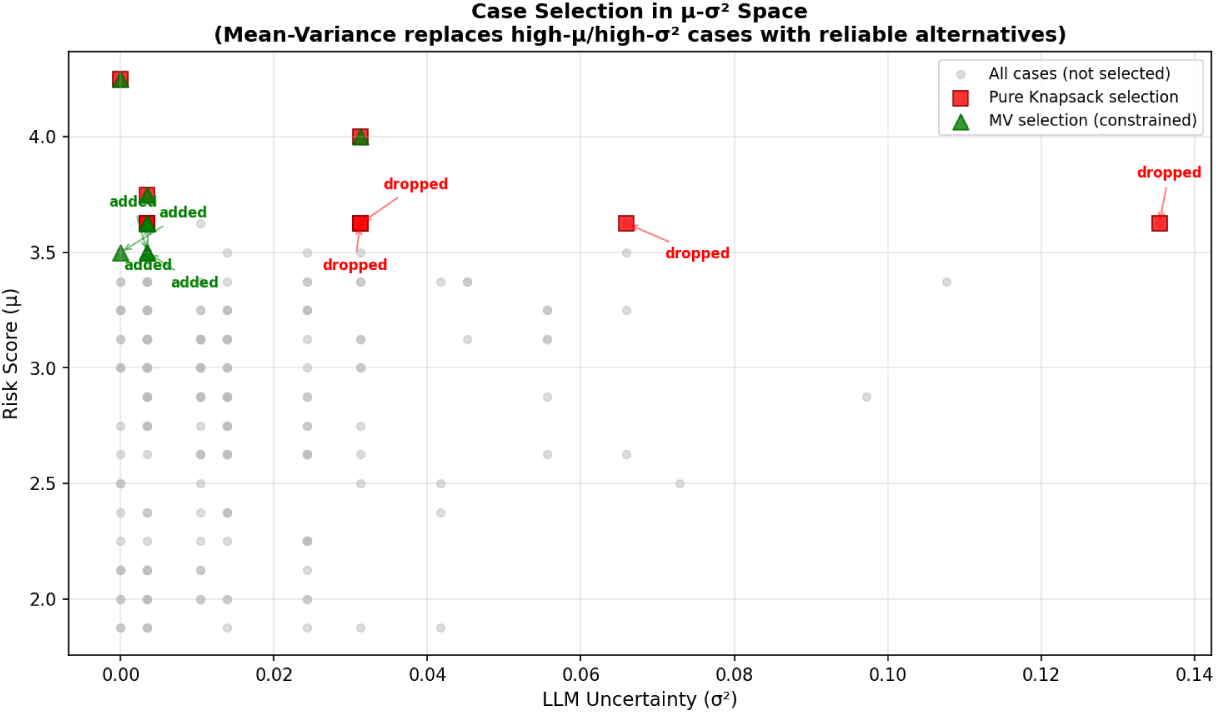
Case selection in *µ*-*σ*^2^ space (synthetic). Red squares = deterministic knapsack selection. Green triangles = mean-variance selection. Annotations show incidents dropped (high-*σ*^2^) and added (low-*σ*^2^) by the mean-variance model.

#### 3.4.4 Replication on Real Cases

Applying the same MV-IP formulation to 196 real TPR cases (*K*=10) revealed a key property of the framework: *the deterministic top-K problem has multiple optimal solutions when ties exist among incidents with equal µ.* An MILP solver without explicit tie-breaking returns one of many optimal-coverage solutions with Σ*µ*=40.750 and Σ*σ*^2^ ranging from 0.205 to 1.274 across 11 distinct coverage-equivalent selections (Figure 4). With explicit tie-breaking via lexicographic ordering by (−*µ, σ*^2^, case id), top-*K* achieves Σ*σ*^2^=0.205. MV-IP, by adding a variance penalty on the equal-coverage solution set, formalizes this as an optimization principle and recovers the same minimum-variance solution (Σ*σ*^2^=0.205). MV-IP therefore does not improve over the best achievable top-*K* solution; rather, it provides a reproducible, optimization-based rule for selecting the most reliably-scored among coverage-equivalent solutions, replacing ad hoc tie-breaking heuristics with an explicit objective. The wide variance range across optimal-coverage selections (0.205 to 1.274) shows that tie-breaking can materially alter selected cases, but it does not establish improved clinical prioritization.

**Fig. 4.**
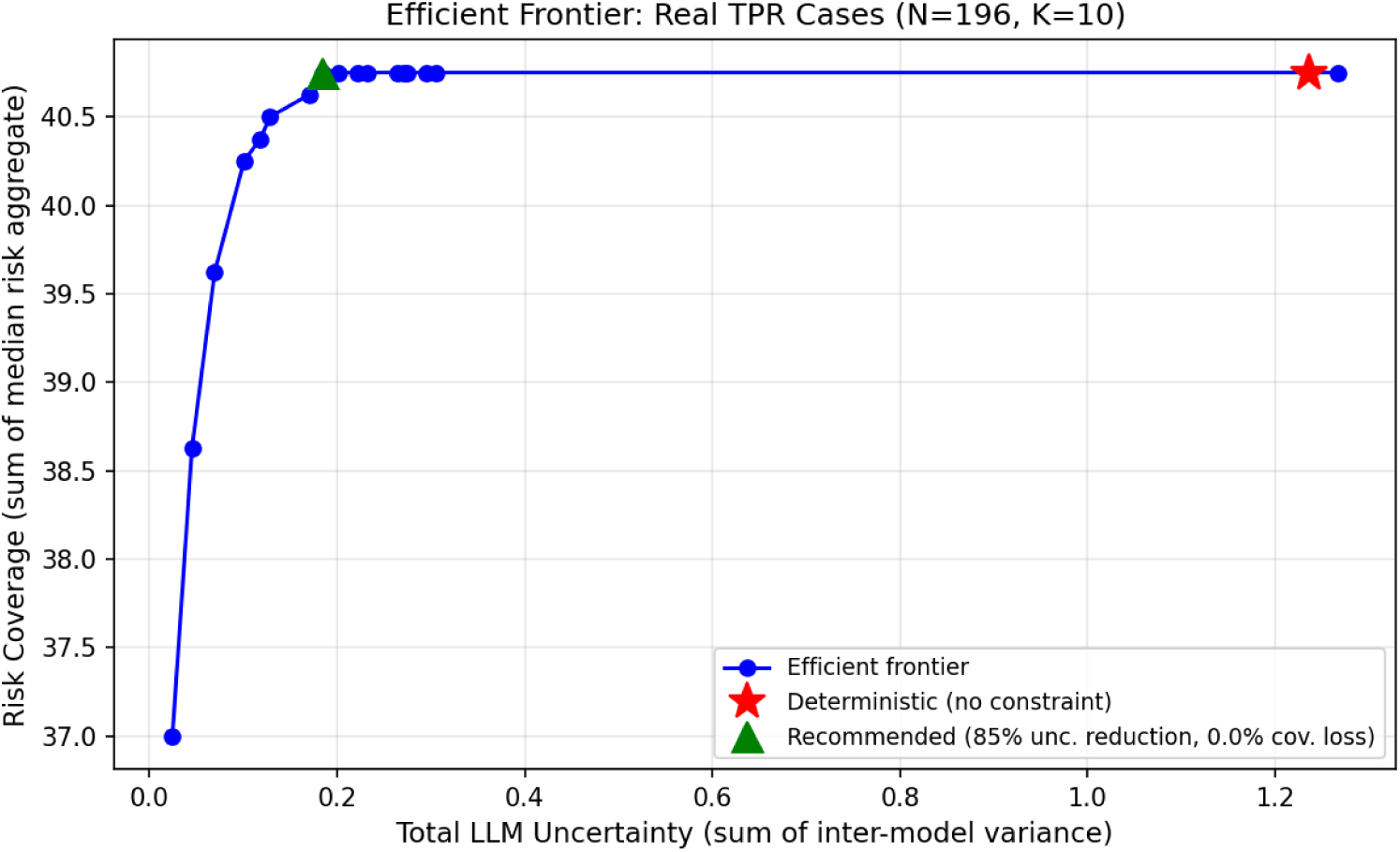
Efficient frontier on real TPR cases (*N* =196, *K*=10). Unlike Figure 2, which sweeps the variance budget to trace a coverage-versus-uncertainty trade-off on synthetic data, this figure shows variance-aware tie-breaking among real-case solutions that all reach the same maximum coverage. At maximum Σ*µ*=40.750, total inter-model variance Σ*σ*^2^ ranges from 0.205 to 1.274 across 11 distinct coverage-equivalent selections.

### 3.5 Sensitivity and Baseline Analyses

#### 3.5.1 Four-Model Robustness Check

To assess whether the 3-model results depend on the specific choice of evaluators, we additionally computed ICC with DeepSeek R1 added as a fourth evaluator (Table 9). On real cases (4-model complete *n*=194 after lab/laboratory category-label normalization), per-dimension ICC ranged 0.256 to 0.829 with changes from the 3-model results within ±0.05 for all dimensions, indicating that the primary findings are robust to evaluator choice. On synthetic cases (4-model complete *n*=199), several dimensions showed larger reductions (D1: 0.903 → 0.794; D2: 0.756 → 0.587), suggesting that synthetic narratives may share generation patterns more readily detected by closely-related model families than by DeepSeek’s distinct reasoning architecture. This further supports the use of real cases as the primary external reliability benchmark.

**Table 9.** Four-model sensitivity check: ICC(A,1) comparison (3-model primary vs 4-model with DeepSeek R1).

| Dimension | Synthetic |  | Real TPR |  |
| --- | --- | --- | --- | --- |
| | 3-model ($n=213$) | 4-model ($n=199$) | 3-model ($n=196$) | 4-model ($n=194$) |
| D1 | 0.903 | 0.783 | 0.834 | 0.829 |
| D2 | 0.756 | 0.587 | 0.565 | 0.573 |
| D3 | 0.832 | 0.832 | 0.699 | 0.720 |
| D4 | 0.783 | 0.704 | 0.529 | 0.489 |
| D5 | 0.539 | 0.449 | 0.275 | 0.256 |
| D6 | 0.868 | 0.796 | 0.745 | 0.731 |
| D7 | 0.693 | 0.661 | 0.593 | 0.615 |
| D8 | 0.892 | 0.786 | 0.782 | 0.776 |

Real-case ICC differences are all within $\pm 0.05$ , supporting robustness. Synthetic ICC reductions on multiple dimensions are interpreted in the discussion.

#### 3.5.2 Test-Retest Reliability (Exploratory)

To assess stochastic noise at temperature=0.1, 30 randomly sampled real cases (seed=42) were re-extracted by each of the four evaluators in a second independent run. Within-model ICC(A,1) between runs ranged 0.482 to 1.000 across all 32 model-dimension combinations (Table 10); 28/32 (87.5%) exceeded 0.70. Gemini achieved perfect reproducibility on D1 (ICC=1.000). DeepSeek showed weaker test-retest on D2 (0.502) and D5 (0.482), reflecting the inherent stochasticity of reasoning-model outputs on judgment-dependent dimensions. As an exploratory analysis with *n*=30 (no category stratification), these results indicate that single-run scores at temperature=0.1 are generally reproducible but should be supplemented by multiple sampling for the most subjective dimensions.

**Table 10.**
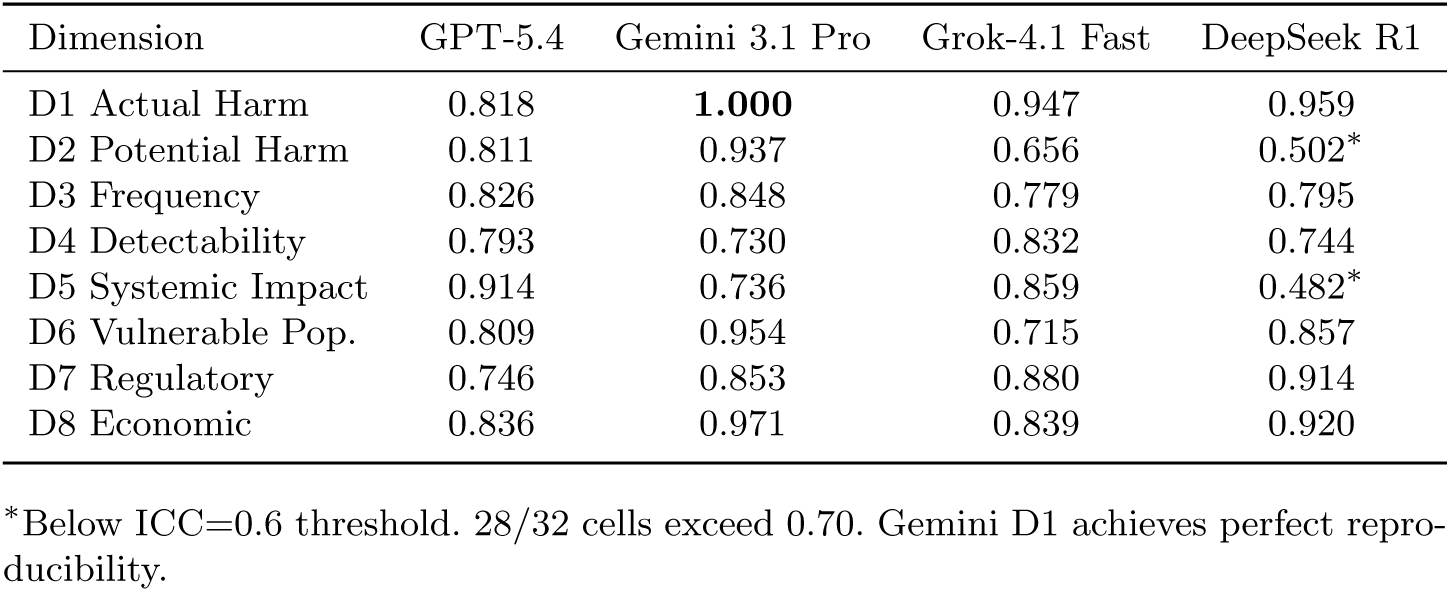
Test-retest within-model ICC(A,1) (random *n*=30 real cases, run 1 vs run 2).

| Dimension | GPT-5.4 | Gemini 3.1 Pro | Grok-4.1 Fast | DeepSeek R1 |
| --- | --- | --- | --- | --- |
| D1 Actual Harm | 0.818 | **1.000** | 0.947 | 0.959 |
| D2 Potential Harm | 0.811 | 0.937 | 0.656 | 0.502* |
| D3 Frequency | 0.826 | 0.848 | 0.779 | 0.795 |
| D4 Detectability | 0.793 | 0.730 | 0.832 | 0.744 |
| D5 Systemic Impact | 0.914 | 0.736 | 0.859 | 0.482* |
| D6 Vulnerable Pop. | 0.809 | 0.954 | 0.715 | 0.857 |
| D7 Regulatory | 0.746 | 0.853 | 0.880 | 0.914 |
| D8 Economic | 0.836 | 0.971 | 0.839 | 0.920 |

\*Below ICC=0.6 threshold. 28/32 cells exceed 0.70. Gemini D1 achieves perfect reproducibility.

#### 3.5.3 External English-Language Narrative Sensitivity Analysis (AHRQ)

As an external English-language narrative sensitivity analysis, we applied the unchanged v1.2 Chinese prompt to 31 English-language AHRQ PSNet WebM&M cases (Table 11). Per-dimension ICC(A,1) point estimates on the English cases were broadly similar to those on the TPR Chinese cases (*n*=196 from Table 8), with within-rater-pair agreement (within-1) ranging 86% to 100%. Three of the four dimensions identified as weakest on Chinese real cases (D2 potential harm Δ=+0.201, D5 systemic impact Δ=+0.196, and D7 regulatory relevance Δ=+0.225) showed higher point estimates on English narratives, but interpretation is limited by sample size (*n*=31) and genre/richness confounding. D4 (detectability) was the only dimension with a slightly lower English ICC (Δ=−0.094), reflecting the wider 95% confidence interval at the smaller English sample. **Interpretation caveat:** this comparison confounds language with document genre and narrative richness: AHRQ WebM&M commentaries provide substantially more explicit discussion of root causes, regulatory implications, and systemic context than TPR brief educational summaries. The higher ICC on D2/D5/D7 may therefore reflect richer textual evidence rather than cross-linguistic transferability per se. These results indicate that the 8-dimensional anchors function on English narratives, though the comparison does not isolate a pure language effect from genre and richness differences.

**Table 11.** External English-language narrative sensitivity analysis: AHRQ English (*n*=31) vs TPR Chinese (*n*=196). Same v1.2 prompt applied; comparison confounds language with document genre and narrative richness.

| Dim | AHRQ English ($n=31$) | | | TPR-ZH | | Note |
| --- | --- | --- | --- | --- | --- | --- |
| | ICC | 95% CI | W-1 | ICC | $\Delta$ | |
| D1 Actual harm | 0.958 | [0.93, 0.98] | 100% | 0.834 | +0.124 | Higher pt. est. |
| D2 Potential harm | **0.766** | [0.62, 0.87] | 98% | 0.565 | **+0.201** | Higher pt. est. |
| D3 Frequency | 0.756 | [0.61, 0.86] | 99% | 0.699 | +0.057 | Comparable |
| D4 Detectability | 0.435 | [0.22, 0.64] | 90% | 0.529 | -0.094 | Lower pt. est. |
| D5 Systemic | **0.471** | [0.25, 0.67] | 86% | 0.275 | **+0.196** | Higher pt. est. |
| D6 Vuln. pop. | 0.769 | [0.63, 0.87] | 88% | 0.745 | +0.024 | Comparable |
| D7 Regulatory | **0.818** | [0.70, 0.90] | 97% | 0.593 | **+0.225** | Higher pt. est. |
| D8 Economic | 0.886 | [0.79, 0.94] | 100% | 0.782 | +0.104 | Higher pt. est. |

#### 3.5.4 Weight Sensitivity

Table 12 shows the impact of four alternative weighting schemes on the MV-IP top-10 selection. Five cases were selected under all four schemes (“robust priority”), demonstrating that the highest-risk incidents are prioritized regardless of weight assumptions. The most divergent scheme (regulatory-heavy) still shared 5 of 10 selections with equal weights (Jaccard = 0.33).

**Table 12.** Sensitivity analysis: weight scheme impact on MV-IP top-10 selection (*N* =213).

| Scheme | Det $\Sigma\mu$ | MV $\Sigma\mu$ | MV $\Sigma\sigma^2$ | Det-MV | Cross |
| --- | --- | --- | --- | --- | --- |
| Equal (1/8 each) | 37.375 | 37.250 | 0.090 | 7/10 | — |
| Harm-heavy (D1,D2=0.2) | 39.400 | 38.960 | 0.071 | 8/10 | 7/10 |
| Regulatory (D7=0.25) | 39.720 | 39.100 | 0.116 | 7/10 | 5/10 |
| Safety-first (D1,2,5,6=0.15) | 37.750 | 37.500 | 0.049 | 8/10 | 8/10 |

Det-MV Overlap = overlap between deterministic and MV-IP within same scheme. Cross-scheme = overlap with Equal-weight MV-IP selection. Five cases were selected under all four schemes regardless of weights.

**Table 13.**
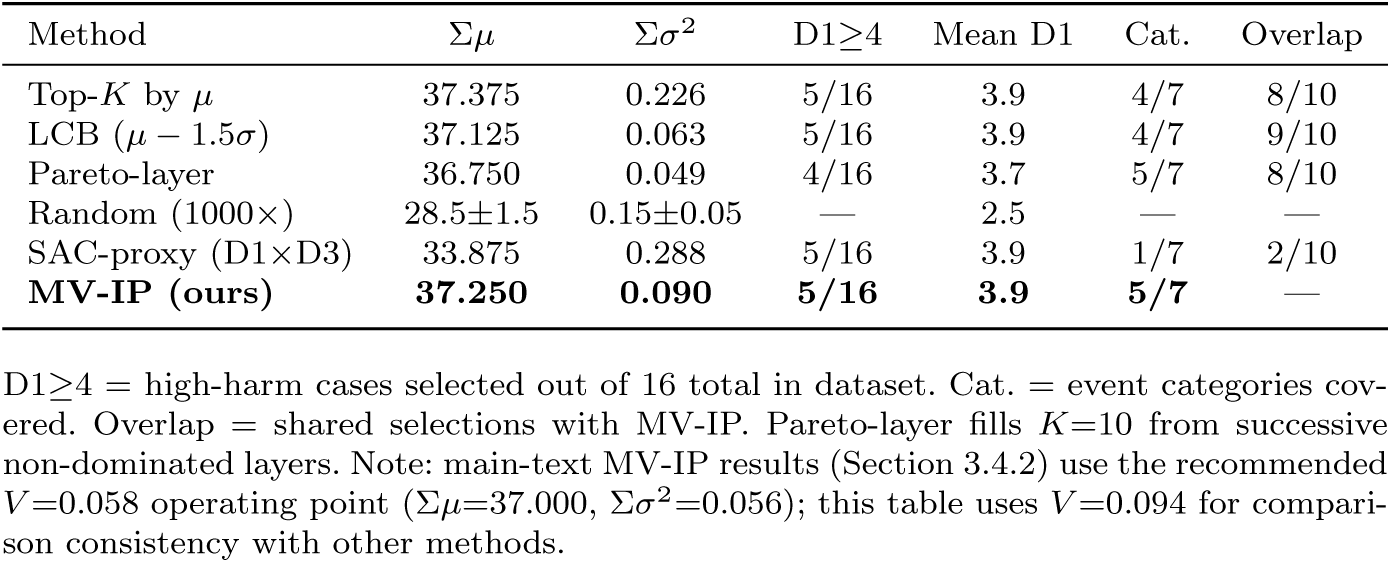
Comparison of six incident selection methods (*K*=10, *N* =213).

The same sensitivity analysis was repeated on real TPR cases (*N* =196). Six cases were selected under all four weighting schemes (vs 5 in synthetic), and Det-MV overlap ranged 8 to 10 out of 10 across schemes (vs 7 to 9 out of 10 in synthetic), confirming that the conclusions about robustness to weight choice generalize to authentic clinical narratives. Full real-case sensitivity results are in supplementary file sensitivity-weights-real.json.

#### 3.5.5 Baseline Comparison

Table 13 compares six selection methods. The MV-IP model achieves the highest category diversity (5/7 categories covered) while maintaining near-optimal coverage, a balance not achieved by any single alternative. The Lower Confidence Bound (LCB) method is the closest alternative, sharing 8/10 selections with MV-IP.

To make the extraction-to-prioritization workflow concrete, Supplementary Table S3 provides an illustrative worked example showing how one synthetic/paraphrased incident narrative is converted into three-model D1 to D8 scores, median aggregate risk, inter-model variance, and MV-IP selection logic. This example is illustrative only and is not included in any reported reliability or optimization metric.

## 4 Discussion

### 4.1 Principal Findings

The central contribution of this work is the extension of patient-safety incident analysis from a two-or three-dimensional paradigm to an eight-dimensional framework, enabled by multi-model LLM extraction and formalized through mean-variance optimization. Three findings merit discussion.

*First*, the 8-dimensional framework successfully operationalizes risk dimensions that quality teams have long considered implicitly but never quantified at scale. By formalizing systemic impact, vulnerable populations, regulatory relevance, and economic consequences alongside the traditional harm-occurrence-detection triad, the framework captures the multi-factorial structure that the RCA community itself recognizes as necessary [7, 8]. The multi-model extraction pipeline demonstrated that three independent LLM families can produce concordant scores on 7 of 8 dimensions (ICC ≥ 0.40 on real cases), establishing that the cognitive bottleneck preventing manual multi-dimensional assessment can be addressed through automation. This direction aligns with recent studies demonstrating the potential of LLMs to scale the analysis of safety reports that currently go unreviewed due to resource constraints [34], and with emerging governance frameworks for generative AI in safety-critical tasks [35].

*Second*, the mean-variance integer optimization provides a principled prioritization mechanism that no prior LLM-FMEA framework has offered. When a deterministic top-*K* solver returns multiple coverage-equivalent solutions (11 distinct selections were observed on real cases, with total inter-model variance ranging 0.205 to 1.274), MV-IP selects the minimum-disagreement set as a reproducible rule. The efficient frontier (Figure 2) makes the coverage-versus-uncertainty trade-off explicit and auditable, offering an extensible framework for incorporating category constraints, capacity constraints, and expert-derived weights via the Best-Worst Method [36]. Clinical validation against expert-prioritized RCA lists is the planned next step.

*Third*, the prompt and model development trajectory suggested that structured reference tables and calibration examples may stabilize extraction. Because prompt version, evaluator model, and sample size changed simultaneously, these observations should guide future controlled ablations rather than be interpreted as causal effects. The model capacity ablation (Supplementary Table S5) is compatible with the possibility that structured prompt design contributed materially to inter-model consistency, while flagship models primarily improved output-format compliance.

Beyond these three findings, two interpretive cautions deserve emphasis. The first concerns the meaning of agreement itself: because no expert ground truth is used here, inter-model agreement is a necessary but not a sufficient condition for a useful score: three models can agree because a dimension is genuinely extractable, or because models trained on overlapping data share the same priors. Our four-model check speaks to this directly. Adding DeepSeek R1, from a different model lineage, left real-case ICC almost unchanged (within 0.05 on every dimension) but reduced several synthetic-case ICCs (D1 from 0.903 to 0.783, D2 from 0.756 to 0.587), consistent with closely related families recognizing shared generation patterns. The same reasoning tempers our high-reliability dimensions: D8 (economic impact) reaches Excellent agreement, yet brief narratives rarely state cost explicitly, so this agreement may reflect a shared severity-to-cost heuristic rather than valid economic estimation. We therefore read high agreement as evidence of reproducibility and explicitly not of clinical correctness, which only the planned expert-consensus study can establish. The second concerns the new dimensions specifically: reliability was not uniform across them. Of the four added beyond the traditional triad, D6 (vulnerable population) and D8 reached Good-to-Excellent agreement on real cases, whereas D5 (systemic impact) was Poor and D7 (regulatory relevance) only Fair, and the single most reliable dimension, D1 (actual harm), is itself a traditional FMEA factor. The contribution is thus better described as identifying which implicitly-used dimensions are already machine-extractable from brief narratives and which are not, rather than as a demonstration that all eight extract reliably; the dimensions that lag (notably D4 and D5) require contextual evidence that brief curated summaries do not contain, which points to a recommendation for incident-reporting form design.

### 4.2 Comparison with Prior Work

El Hassani et al. [16] pioneered LLM integration with FMEA but used a single model family (GPT) for both generation and evaluation, retained the 3-dimensional RPN framework, and provided no external reliability evaluation (Table 14). Our work differs in design through strict model separation across four provider families [17], extension to 8 dimensions grounded in established quality frameworks [21, 23], dual-dataset reliability assessment including 196 curated educational TPR cases, and quantified inter-model consistency via ICC [25]. Direct head-to-head metric comparison is limited by domain differences and the absence of shared benchmark data; the table therefore summarizes design-level rather than performance-level comparison.

**Table 14.**
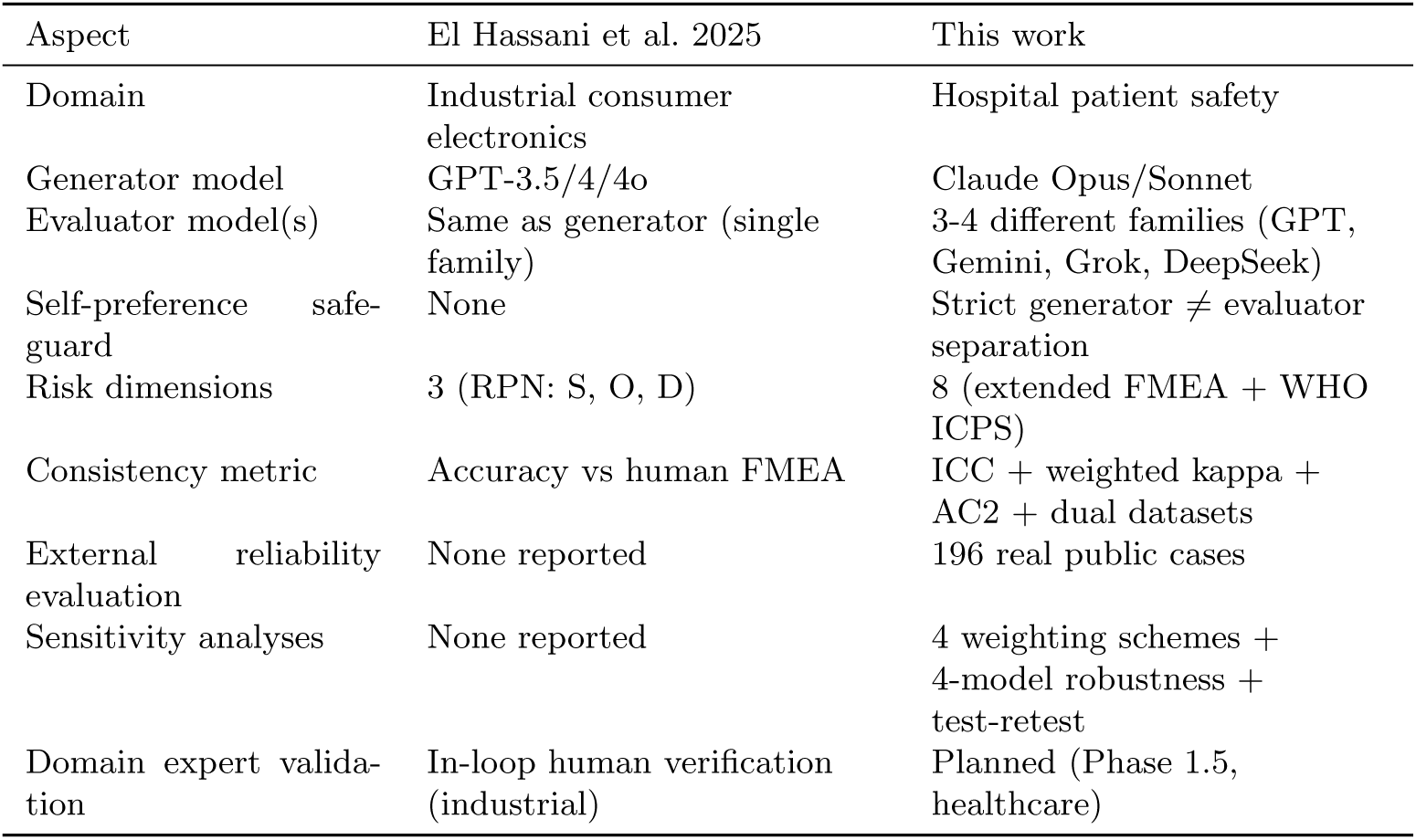
Design-level comparison with El Hassani et al. (2025).

Within the broader FMEA improvement literature [4], our work touches three of the five improvement directions identified in systematic reviews: dimension extension (D5 to D8), AI integration (multi-model LLM), and a prototype treatment of uncertainty (inter-model variance *σ*^2^ as an explicit parameter). Where prior approaches typically pursued these directions one at a time, the present framework brings them together; we note, however, that the dimension extension is reliably extractable only for some of the new dimensions and that the uncertainty layer is a formalization rather than demonstrated evidence of improved prioritization.

The mean-variance optimization adds a secondary uncertainty-management layer absent from prior LLM-FMEA work. By treating *σ*^2^ (inter-model variance) as explicit extraction uncertainty, it bridges NLP extraction and operations research while remaining a prototype formalization rather than evidence of improved clinical prioritization.

### 4.3 Relation to Classical Decision Matrices and Commercial RCA Systems

The 8-dimensional risk feature framework can be reinterpreted within the long lineage of decision matrices in quality management and industrial engineering. Classical Quality Control Circle (QCC) practice routinely employs a Solution Evaluation Matrix in which 5 to 10 candidate countermeasures are scored across 4 to 5 criteria (effectiveness, feasibility, economy, time, autonomy) by team consensus, weighted, summed, and compared to an adoption threshold. Pugh’s concept selection matrix [37], the Analytic Hierarchy Process (AHP) [38], TOPSIS [18], and the Best-Worst Method (BWM) [36] are well-established generalizations of this paradigm. Our framework can be viewed as a scalable, uncertainty-aware extension of this lineage: where a QCC team scores 5 alternatives × 5 criteria via consensus, our pipeline scores 200+ incidents × 8 dimensions via three independent LLMs, with each cell carrying an explicit between-rater variance. The mean-variance integer program extends “argmax row-sum subject to threshold” by making model disagreement explicit when many alternatives share equivalent coverage. Three structural differences merit emphasis: (a) *direction*: classical decision matrices choose among future actions, whereas our framework prioritizes past events for retrospective investigation; (b) *scale*: automation may reduce the operational burden that historically limited matrices to 5 to 10 rows and 4 to 5 columns; and (c) *uncertainty*: inter-rater variance, which classical matrices treated only informally via discussion, becomes an explicit optimization variable.

Within the patient-safety domain specifically, two reference points warrant explicit positioning. The AHRQ Patient Safety Network’s WebM&M (Morbidity & Mortality Rounds on the Web), running since 2003 [32], has accumulated hundreds of de-identified incident cases with peer-reviewed expert commentary, providing a public corpus suitable for external sensitivity analysis (as we demonstrate in Section 3.5.3). Commercial RCA systems such as TapRooT^®^ [39], accepted by The Joint Commission and adopted by hospital quality programs, codify root-cause investigation through a proprietary 7-category Root Cause Tree^®^ Dictionary and a 15-question Human Performance Troubleshooting Guide, walking investigators through structured causal categorization of an already-identified event. Our framework occupies a complementary, upstream role: rather than guiding within-event causal analysis, it operationalizes a multi-dimensional triage at the *between-event prioritization* stage that determines which incidents enter formal RCA at all. The two paradigms can coexist, with LLM-extracted 8-dimensional triage selecting which incidents warrant TapRooT-style or HFMEA-style structured analysis, rather than competing.

### 4.4 Prompt and Model Development Observations

The model capacity ablation (flagship vs. lighter models) revealed near-identical pooled ICC (0.713 vs. 0.716), while parse success rate improved from 58% to 98% complete-case extraction (Supplementary Table S5). This pattern is compatible with the possibility that structured prompts and calibration examples contributed materially to inter-model consistency, while flagship models primarily improved output-format compliance, consistent with recent evidence on prompt engineering methodology [14, 40]. The D3 improvement (ICC 0.567 → 0.832) coincided with both a frequency reference table addition and a model version upgrade; controlled ablation is needed to isolate prompt effects.

### 4.5 D5 (Systemic Impact): A Theoretical Tension

The framework’s weakest dimension on real data, D5 (Systemic Impact, ICC=0.275), is also its most theoretically motivated, anchored in Reason’s Swiss Cheese Model and Vincent’s London Protocol. Detailed examination reveals a specific quantitative pattern that is mathematically distinct from low rater agreement (Supplementary Table S2). Across 196 real cases, 73.5% received identical D5 scores from all three evaluators, and the score distribution clusters tightly around the modal value of 3 (74% of GPT scores, 84% of Gemini, 90% of Grok). The between-case standard deviation of D5 score medians on real cases is only 0.41, compared with 0.97 for D1 (actual harm) on the same dataset, a 2.4-fold reduction in between-case variance. ICC mathematically depends on between-case variance (*σ*^2^cases/[*σ*^2^cases+*σ*^2^error]); when *σ*^2^cases approaches zero, ICC approaches zero regardless of error magnitude. When evaluators disagreed on D5, 95% of disagreements were within ±1 (only 11 of 196 cases showed differences ≥2 between any two models). This pattern suggests that poor ICC reflects both compressed between-case variance and limited textual discrimination in brief narratives, rather than frequent large-magnitude model disagreement.

This is consistent with the restricted-range explanation [26, 30]: low ICC arises mathematically when between-case variance approaches zero, regardless of within-case agreement. However, the more important clinical interpretation is that D5 may not be reliably discriminative from brief curated narratives. The available text may simply lack enough contextual evidence for LLMs to distinguish individual errors from system-level failure. D5 is therefore interpreted as theoretically important but methodologically under-supported by brief narratives. Future work should separate explicit systemic-factor detection from severity grading (e.g., D5a: systemic factor present/absent; D5b: systemic impact severity when evidence is present) and require evidence spans supporting high D5 scores. Distinguishing true homogeneity in reporting culture from insufficient textual evidence requires expert validation, which is outside the present study’s scope.

### 4.6 Generalizability: Synthetic to Real

The ICC reduction from synthetic to real cases reflects the greater clinical ambiguity in real narratives: longer texts, more nuanced descriptions, and information distributed across educational commentary rather than structured fields. Importantly, within-1 agreement remained ≥72% across all 8 dimensions on real cases, indicating that most inter-model differences were within one ordinal score category. This should be interpreted as a reliability pattern, not evidence that the scores are clinically acceptable or clinically valid.

The lower reliability of D4, D5, D7, and D8 on real cases suggests that not all risk dimensions are equally extractable from brief incident narratives. These dimensions may require structured reporting fields (e.g., explicit detectability descriptions) or expert adjudication rather than fully automated extraction, a finding with implications for incident reporting form design.

### 4.7 Multi-Model Panel Composition

The four-model robustness check (Table 9) showed that leave-one-out ICC changes remained within ±0.05 on real cases for all dimensions, indicating that no single evaluator dominates the primary 3-model results. At the same time, the MV-IP leave-one-out analysis revealed selection Jaccard indices of 0.43 to 0.67 across evaluator-drop configurations, indicating that each model contributes non-redundant scoring perspective to the top-*K* prioritization outcome. This pattern supports the multi-model design rationale: a single-model panel would lack the inter-model variance signal that MV-IP requires, while the observed non-redundancy confirms that the three evaluators capture genuinely diverse scoring perspectives rather than producing redundant copies of the same pattern. These findings caution against single-model deployment for prioritization tasks where model disagreement serves as an uncertainty signal.

### 4.8 Alignment with International and Local Patient Safety Priorities

Approximately 44% of incidents in national patient safety reporting systems are attributed to human factors, rising to nearly 60% when communication-related factors are included [24]. This dominance of human-and-system-level pathways underscores why a framework restricted to harm severity and occurrence frequency (traditional RPN) cannot capture the structural risk that drives many incidents. The framework’s D5 (systemic impact) dimension directly targets this pathway conceptually, but the present reliability results show that brief curated narratives may not provide enough evidence for automated discriminative scoring of systemic impact.

The World Health Organization’s Global Patient Safety Action Plan 2021 to 2030 explicitly designates Human Factors Engineering (HFE) as a core strategy for high-reliability health systems and identifies the International Ergonomics Association as a key implementation partner [41]. Notably, the VA National Center for Patient Safety has long observed that “Human Factors Engineering actions work best, but training, writing policies, and reminders to pay more attention are generally ineffective” [1]. Within this policy direction, automated multi-dimensional risk extraction can serve a specific HFE-aligned function: identifying incidents whose D4 (low detectability) and D5 (high systemic impact) profiles indicate that further training or policy revision is unlikely to suffice and that engineering-level interventions (forcing functions, redesigned interfaces, or barrier systems) are required.

### 4.9 Future Dual-Track Triage Concept

The efficient frontier illustrates a conceptual trade-off, not a validated operational tool. An important clinical consideration arises: in patient safety, should incidents with high risk scores but high LLM disagreement be deprioritized, or should they be escalated for human review? Unlike financial portfolios where high-variance assets are simply avoided, high-uncertainty incidents in healthcare may represent precisely the cases requiring careful human judgment.

A dual-track operational framework is proposed:

- **RCA Priority Queue** (high *µ*, low *σ*^2^): Incidents with high risk scores and strong inter-model agreement. In a future validated workflow, these would be candidates for direct RCA review.
- **Adjudication Queue** (high *µ*, high *σ*^2^): Incidents with high risk scores but substantial inter-model disagreement. Rather than being excluded by optimization, these would be flagged for human review in triage meetings, where domain experts resolve the ambiguity.

At the recommended operating point, the 4 incidents replaced by the mean-variance model represent candidates for the adjudication queue rather than incidents to be ignored. This reframing aligns the optimization with clinical practice, where uncertainty triggers escalation rather than dismissal.

If validated against expert consensus in Phase 1.5, the framework could support, though not replace, manual review of 50 to 70 monthly incident reports by providing automated multi-dimensional risk profiling as a structured pre-screening layer.

### 4.10 Limitations

Three limitations merit consideration. *First*, clinical validity against expert consensus has not yet been established. Inter-model concordance demonstrates extraction reproducibility, not clinical correctness; a Phase 1.5 expert panel study is in preparation to assess calibration against clinician-prioritized RCA lists. *Second*, D5 (Systemic Impact) showed poor discriminative reliability (ICC = 0.275) despite high within-1 agreement, reflecting compressed score variance from brief narrative inputs rather than frequent large-magnitude disagreement (Section 4.5); richer narrative formats or explicit systemic-factor reporting fields may improve discrimination. *Third*, the vari-ance parameter *σ*^2^ is estimated from *k*=3 evaluators (df=2); bootstrap analysis showed that 74% to 90% of per-case estimates have narrow 95% CIs, but cases with wide CIs represent selection-sensitive inputs to MV-IP. Expanding the evaluator panel and exploring factor-model variance estimators are natural extensions.

## 5 Conclusions

This study makes three contributions to patient-safety incident analysis.

*First*, the 8-dimensional risk feature framework extends the traditional 3-dimensional FMEA (Severity × Occurrence × Detection) to capture systemic impact, vulnerable populations, regulatory relevance, and economic consequences, dimensions that quality teams consider implicitly but have not previously operationalized at scale. The framework formalizes root cause analysis triggering logic into machine-extractable Likert scales, enabling structured risk profiling across hundreds of incident narratives.

*Second*, the multi-model LLM extraction pipeline demonstrates that three independent evaluator families can produce concordant 8-dimensional risk scores from unstructured incident narratives. On 196 real cases spanning seven event categories, seven of eight dimensions reached Fair or better inter-model reliability (ICC at least 0.40), though reliability was not uniform: two dimensions were Excellent and three only Fair, so some dimensions are more readily extractable than others. The framework additionally functioned on English-language narratives. D5 (Systemic Impact) is retained as theoretically motivated but requires richer narrative input for reliable discrimination.

*Third*, the variance-aware integer optimization (MV-IP) formalizes tie-breaking under extraction uncertainty. It does not increase risk coverage over a deterministic top-*K* solver; rather, when several selections share the same coverage, it treats inter-model variance as an explicit parameter and selects the most reliably scored set, replacing ad hoc tie-breaking with a reproducible rule. Because low disagreement is used only to choose among equal-coverage sets, it is not a claim that contentious incidents matter less: the dual-track design routes high-confidence incidents to RCA review and flags high-disagreement incidents for expert adjudication, so that uncertainty triggers escalation rather than exclusion. This workflow is intended to support, not replace, a future expert-validated process.

Expert-consensus validation against clinician-prioritized RCA lists (Phase 1.5), institutional deployment on raw incident reports (Phase 2), and expert-derived dimension weights via the Best-Worst Method [36] are the planned next steps toward clinical translation. This study constitutes one component of an umbrella research programme on LLM-assisted hospital quality improvement, within which methodological extensions of the present optimization framework, incorporating additional outcome dimensions beyond risk such as resource and sustainability considerations, are being developed as separate companion studies.

## Abbreviations

*AC2*: Gwet’s agreement coefficient
*AHRQ*: Agency for Healthcare Research and Quality
*BWM*: Best-Worst Method
*FMEA*: Failure Mode and Effects Analysis
*HFMEA*: Healthcare Failure Mode and Effects Analysis
*ICC*: intraclass correlation coefficient
*ICPS*: International Classification for Patient Safety
*LLM*: large language model
*MILP*: mixed-integer linear programming
*MV-IP*: mean-variance integer programming
*PSNet*: Patient Safety Network
*RCA*: root cause analysis
*RPN*: Risk Priority Number
*SAC*: Severity Assessment Code
*TPR*: Patient-safety Reporting system

## Supplementary information

Supplementary information accompanies this paper and includes four files: Supplementary File S1 (extraction prompt and scoring rubric), Supplementary File S2 (MV-IP formulation and worked example), Supplementary File S3 (sensitivity analysis on AHRQ PSNet cases), and Supplementary File S4 (data provenance and case-source registry).

## Data Availability

All data produced in the present study are available upon reasonable request to the authors

## Acknowledgements

The authors thank the patient-safety reporting and quality-improvement communities whose publicly available educational materials made this methodological reliability study possible. No generative AI system was used to make clinical judgments or to replace expert review in this study; all LLM outputs were treated as algorithmic extractions for reliability analysis.

## Author contribution

HML conceived the study, designed the 8-dimensional framework, developed the LLM extraction workflow, conducted the analyses, interpreted the results, and drafted the manuscript. JL and ILW provided methodological supervision, industrial and information-management expertise, and critical review of the framework, optimization formulation, and manuscript. All authors reviewed and approved the final manuscript.

## Data availability

The datasets generated and analysed during the current study, including the complete extraction prompt (v1.2), system prompt, parse rules, model and run provenance appendix, analysis code, and nonidentifiable derived outputs, are available from the corresponding author on reasonable request. Public TPR and AHRQ PSNet source materials remain subject to the terms of their original publishers. No identifiable patient-level institutional incident reports are included in the submitted materials.

## Code availability

The extraction pipeline, analysis scripts, and supplementary code are available from the corresponding author on reasonable request. Commercial LLM API access is required for full pipeline reproduction.

## Declarations

### Ethics approval and consent to participate

Institutional review board approval and participant consent were not required because this methodological reliability study used LLM-generated synthetic cases and publicly available, curated educational patient-safety narratives. No identifiable patient-level institutional incident reports were used.

### Consent for publication

Not applicable.

### Relevant guidelines and regulations

All methods were performed in accordance with relevant guidelines and regulations for studies not involving human subjects. No identifiable patient data were used in this study.

