## supplementary for "Operationalizing Eight-Dimensional Patient-Safety Risk Scoring at Scale: A Multi-Model Large Language Model Reliability Study"

#### Supplementary Table S1. Case Attrition by Stage and Category

##### Dataset A: Synthetic Incident Cases

Table 1: Dataset A case attrition by processing stage.

| Stage | N | Attrition | Cumulative retained |
|---|---|---|---|
| Generated target | 300 | Reference stage | 100% |
| Loaded, YAML parsed successfully | 255 | 45 loading failures | 85.0% |
| Complete extraction, all 3 evaluators x 8 dimensions | 213 | 42 parse failures | 71.0% |

Target generation distribution was medication 30%, fall 15%, tube/line 10%, surgery 10%, laboratory 10%, transfusion 5%, and other 20%.

Table 2: Dataset A loaded cases by category.

| Category | Target N | Loaded N | Loading attrition |
|---|---|---|---|
| Medication | 90 | 79 | 11 |
| Fall | 45 | 37 | 8 |
| Tube/line | 30 | 26 | 4 |
| Surgery | 30 | 27 | 3 |
| Laboratory | 30 | 27 | 3 |
| Transfusion | 15 | 11 | 4 |
| Other | 60 | 48 | 12 |
| Total | 300 | 255 | 45 |

Loaded category counts were reconstructed from the successfully loaded synthetic case source files and cross-checked against the 255-case primary-attempted manifest. Loading failures (N=45) were caused by malformed YAML output from the generator LLM before evaluator extraction. A goodness-of-fit chi-squared test comparing the 255 successfully loaded cases against the 300-case target generation proportions was not significant ($\chi^2$=0.73, df=6, p=0.994), indicating that loading failures were not systematically associated with event category.

Parse failures (N=42) were caused by at least one evaluator returning output that could not be parsed into valid 8-dimensional scores. Per-model parse failure rates among the 255 loaded cases were GPT-5.4 21/255 (8.2%), Gemini 3.1 Pro 41/255 (16.1%), and Grok-4.1 Fast 0/255 (0.0%). Category-specific extraction failure counts could not be formally assessed because extraction output files do not contain event category labels.

#### Dataset B: Real TPR Cases

Table 3: Dataset B real TPR case attrition by category.

| Category | Available | Complete extraction | Attrition |
|---|---|---|---|
| Medication | 68 (34.3%) | 67 (34.2%) | 1 |
| Other | 61 (30.8%) | 61 (31.1%) | 0 |
| Tube/line | 23 (11.6%) | 23 (11.7%) | 0 |
| Surgery | 22 (11.1%) | 22 (11.2%) | 0 |
| Fall | 11 (5.6%) | 11 (5.6%) | 0 |
| Laboratory | 10 (5.1%) | 9 (4.6%) | 1 |
| Transfusion | 3 (1.5%) | 3 (1.5%) | 0 |
| Total | 198 (100%) | 196 (100%) | 2 |

Table 4: Excluded real TPR cases.

| Case ID | Category | Failure | Cause |
|---|---|---|---|
| NO.113 | Medication | Evaluator 3, Grok-4.1 Fast | Incomplete or malformed YAML output with missing dimensions |
| NO.175 | Laboratory | Evaluator 3, Grok-4.1 Fast | Incomplete or malformed YAML output with missing dimensions |

Both real-case exclusions were caused by Grok-4.1 Fast returning structurally invalid YAML. GPT-5.4 and Gemini 3.1 Pro returned complete scores for both cases. No category proportion changed by more than 0.5 percentage points.

#### Supplementary Table S2. D5 Systemic Impact Diagnostics

This supplement expands the main-text interpretation of D5 (Systemic Impact), the only dimension with Poor ICC on real TPR cases. The purpose is to distinguish low ICC caused by restricted between-case variance from large model disagreement.

Table 5: D5 agreement diagnostics by dataset.

| Dataset | N | ICC(A,1) | Within-1 | Exact agreement | Interpretation |
|---|---|---|---|---|---|
| Synthetic cases | 213 | 0.539 | 97% | 88.1% | Fair ICC with strong absolute agreement; restricted marginals visible. |
| Real TPR cases | 196 | 0.275 | 95% | 73.5% | Poor ICC despite high within-1 agreement; D5 is not reliably discriminative from brief curated narratives. |
| AHRQ English narratives | 31 | 0.471 | 86% | Not estimated | Fair ICC, likely supported by richer narrative context and expert commentary format. |

D5 is retained because systemic impact is central to patient-safety theory and RCA practice, but it should not be interpreted as deployment-ready for fully automated discrimination from brief curated narratives. The observed pattern supports two simultaneous conclusions: models often agree closely on D5 scores, and the brief TPR narrative format may not contain enough contextual evidence to distinguish individual error from system-level failure. Future validation should separate explicit systemic factor detection from severity grading and should require evidence spans for high D5 scores.

Table 6: Real TPR D5 restricted-range pattern.

| Diagnostic | Value | Reviewer-facing interpretation |
|---|---|---|
| Real-case D5 ICC(A,1) | 0.275 | Poor by Cicchetti benchmark. |
| Real-case D5 within-1 agreement | 95% | Most disagreements are within one ordinal category. |
| Cases with identical D5 score from all three evaluators | 73.5% | High exact agreement despite low ICC. |
| Cases with any pairwise D5 difference $\geq 2$ | 11/196 | Large model disagreement is uncommon. |
| Between-case SD of D5 median score | 0.41 | Very limited between-case variation suppresses ICC. |
| Between-case SD of D1 median score | 0.97 | D5 has 2.4x lower between-case spread than actual harm. |

#### Supplementary Table S3. Worked Example of the 8-Dimensional Extraction and MV-IP Pipeline

This worked example is illustrative and uses a synthetic or paraphrased medication incident to avoid patient privacy and copyright concerns. It is not used as additional empirical evidence and does not change any reported study results.

##### Incident Narrative

A hospitalized older adult with chronic kidney disease was prescribed a renally cleared antibiotic. The initial dose was entered using the standard adult dose. Pharmacy verification did not flag the renal-dose issue because the estimated glomerular filtration rate field was not visible in the medication verification screen. The patient developed transient confusion and nausea after two doses. The medication was held after nursing staff contacted the physician. The patient recovered without intensive care transfer. The hospital later identified that renal dosing alerts were inconsistently configured across medication classes.

Table 7: Three-model 8-dimensional scores.

| Dim. | Meaning | GPT | Gemini | Grok | Median | Range |
|---|---|---|---|---|---|---|
| D1 | Actual harm | 3 | 3 | 3 | 3 | 0 |
| D2 | Potential harm | 4 | 4 | 5 | 4 | 1 |
| D3 | Base frequency | 3 | 3 | 3 | 3 | 0 |
| D4 | Detectability | 4 | 3 | 4 | 4 | 1 |
| D5 | Systemic impact | 4 | 3 | 4 | 4 | 1 |
| D6 | Vulnerable population | 4 | 4 | 4 | 4 | 0 |
| D7 | Regulatory relevance | 3 | 3 | 2 | 3 | 1 |
| D8 | Economic impact | 2 | 2 | 3 | 2 | 1 |

Table 8: Aggregate risk and uncertainty in the worked example.

| Model or aggregate | Mean of 8 dimensions |
|---|---|
| GPT-5.4 | 3.375 |
| Gemini 3.1 Pro | 3.125 |
| Grok-4.1 Fast | 3.500 |
| Median aggregate risk, $\mu$ | 3.375 |
| Inter-model variance, $\sigma^2$ | 0.036 |

This case would be interpreted as moderate-to-high priority because the median aggregate risk is elevated and model disagreement is modest. Clinically, D2, D4, D5, and D6 drive the priority profile: the event involved a vulnerable patient, nontrivial potential harm, imperfect detectability, and possible system-level decision-support gaps. Under deterministic top-K selection, the case would be ranked by $\mu$. Under MV-IP, the case would remain attractive if it lies on the equal-coverage frontier because its disagreement variance is low relative to other cases with similar aggregate risk.

### Supplementary File S4. Provenance Appendix

#### Study Metadata

Table 9: Study metadata.

| Item | Value |
|---|---|
| Study | Q02 / Sub-study 2 |
| Design | Multi-model reliability study with secondary uncertainty-aware prioritization prototype |
| Primary extraction framework | 8-dimensional patient-safety risk feature extraction |
| Primary synthetic set | 300 generated; 255 primary-attempted; 213 complete 3-model extractions |
| Primary real TPR set | 198 design cases; 196 complete 3-model extractions |
| English-language sensitivity set | 31 AHRQ PSNet WebM&M narratives |
| Primary prompt | `feature-extraction-v1.2.md` |
| Prompt version | v1.2 |
| Main analysis manifest | `ACTIVE_RESULTS_MANIFEST.md` |
| Manuscript package | `Q02_BMC-MIDM_20260513/` |

Table 10: Model provenance.

| Dataset/run | Eval. | Provider model ID | Route | Cap | Analysis role |
|---|---|---|---|---|---|
| Synthetic primary | GPT | `openai/gpt-5.4` | OpenRouter OpenAI-compatible route | 2048 | Primary 3-model evaluator |
| Synthetic primary | Gemini | `gemini-3.1-pro-preview` | Google GenAI SDK | 8192 | Primary 3-model evaluator |
| Synthetic primary | Grok | `x-ai/grok-4.1-fast` | OpenRouter route | 2048 | Primary 3-model evaluator |
| Real TPR primary | GPT | `gpt-5.4` | OpenAI API | 2048 | Primary 3-model evaluator |
| Real TPR primary | Gemini | `gemini-3.1-pro-preview` | Google GenAI SDK | 8192 | Primary 3-model evaluator |
| Real TPR primary | Grok | `x-ai/grok-4.1-fast` | OpenRouter route | 2048 | Primary 3-model evaluator |
| AHRQ sensitivity | GPT | `gpt-5.4` | OpenAI API | 2048 | English-language narrative sensitivity |
| AHRQ sensitivity | Gemini | `gemini-3.1-pro-preview` | Google GenAI SDK | 8192 | English-language narrative sensitivity |
| AHRQ sensitivity | Grok | `x-ai/grok-4.1-fast` | OpenRouter route | 2048 | English-language narrative sensitivity |
| 4-model/retest sensitivity | DeepSeek | `deepseek/deepseek-r1` | OpenRouter route | 8192 | 4-model robustness and retest extension |

#### Run Configuration

#### Dataset, Extraction, and Analysis Artifacts

Primary scripts documenting model identifiers are under `04-analysis/sub-study-2/phase1-notebooks/`: `run_feature_extraction.py`, `run_tpr_extraction_strong.py`, `run_tpr_new143_extraction.py`, `run_deepseek_all.py`, `run_ahrq_extraction.py`, and `run_test_retest.py`.

Table 11: Run configuration.

| Item | Value |
|---|---|
| Temperature | 0.1 |
| OpenRouter base URL | `https://openrouter.ai/api/v1` when OpenRouter was used |
| OpenAI-compatible max output budget | 2048 tokens for GPT/Grok primary extraction calls |
| Gemini SDK max output budget | 8192 tokens in scripts using Google GenAI SDK |
| DeepSeek max output budget | 8192 tokens in selected DeepSeek/test-retest calls |
| Prompt delivery | System prompt loaded from `feature-extraction-v1.2.md`; user prompt wraps each incident narrative |
| Retry/repair policy | No hidden manual score correction; parsed output required valid 8-dimensional scores |
| Complete-case policy | Primary ICC analyses use cases with complete valid scores across the relevant evaluator set |

Table 12: Analysis artifacts and manuscript roles.

| Artifact | Manuscript role |
|---|---|
| `4-model-icc-results.json` | 4-model robustness |
| `agreement-supplementary.json` | Supplementary agreement metrics |
| `parse-failure-analysis.json` | Pipeline attrition and parse-failure analysis |
| `d5-error-analysis.json` | D5 restricted-range and agreement diagnostics |
| `test-retest-icc.json` | Test-retest reliability |
| `sensitivity-weights-results.json` | Synthetic weight sensitivity |
| `sensitivity-weights-real.json` | Real-case weight sensitivity |
| `module-b-mv-results.json` | Synthetic MV-IP frontier |
| `baseline-comparison-real.json` | Real-case baseline comparison |

#### Parse and Complete-Case Policy

**Table S4.3. Parse and complete-case policy.**

| Dataset | Design/attempted | Complete 3-model | Complete 4-model | Notes |
|---|---|---|---|---|
| Synthetic | 300 generated; 255 attempted | 213 | 199 | YAML/load failures and model parse failures are pipeline reliability endpoints. |
| Real TPR | 198 design cases | 196 | 194 | TPR category labels are normalized for lab/laboratory variants. |
| AHRQ sensitivity | 31 public English cases | 31 | Not primary | Interpreted as narrative sensitivity, not isolated language transfer. |

#### Software Environment Snapshot

**Table S4.4. Software environment snapshot captured on 2026-05-10.**

| Package | Version |
|---|---|
| Python | 3.13.9 |
| pandas | 2.3.3 |
| numpy | 2.4.0 |
| scipy | 1.17.1 |
| statsmodels | 0.14.6 |
| requests | 2.32.5 |
| openai | 2.32.0 |
| python-docx | available in local environment |

##### Repository Snapshot and Provenance Gaps

**Table S4.5. Repository snapshot.**

| Item | Value |
|---|---|
| Git commit at provenance capture | `e6c79e8` |
| Workspace state | Dirty at capture time |
| Active manuscript root | `Q02_BMC-MIDM_20260513/` |
| LaTeX source package | `Q02_BMC-MIDM_20260513/latex/` |
| Supplementary source package | `Q02_BMC-MIDM_20260513/supplementary/` |

Remaining provenance gaps include incomplete provider-side request timestamp export, unavailable provider-side served model release hashes, partially centralized token accounting, and external public-source versioning for TPR/AHRQ pages. These gaps are disclosed as provenance limitations rather than hidden assumptions.

#### Supplementary Table S5. Model Capacity Ablation: Per-Dimension ICC

| | ICC(A,1) | | | |
|---|---|---|---|---|
| Dimension | Lighter ($n$=32) | Flagship ($n$=54) | Δ | Direction |
| D1 Actual Harm | 0.729 | 0.702 | −0.027 | Lighter higher |
| D2 Potential Harm | 0.647 | 0.784 | +0.137 | Flagship higher |
| D3 Frequency | 0.553 | 0.621 | +0.068 | Flagship higher |
| D4 Detectability | 0.627 | 0.500 | −0.127 | Lighter higher |
| D5 Systemic Impact | 0.100 | 0.212 | +0.112 | Flagship higher |
| D6 Vulnerable Pop. | 0.723 | 0.811 | +0.088 | Flagship higher |
| D7 Regulatory | 0.424 | 0.536 | +0.112 | Flagship higher |
| D8 Economic Impact | 0.504 | 0.526 | +0.022 | Flagship higher |
| Mean per-dimension | 0.538 | 0.587 | +0.049 | Flagship higher |
| Pooled (all 8 dims) | 0.716 | 0.713 | −0.003 | Near-identical |

Lighter panel: GPT-4.1, Gemini 2.5 Flash, Grok-3-mini. Flagship panel: GPT-5.4, Gemini 3.1 Pro, Grok-4.1 Fast. Same v1.2 prompt applied to 55 real TPR cases. Complete-case $n$ differs because lighter models had higher parse failure rates (lighter complete: 32/55 = 58%; flagship complete: 54/55 = 98%). Pooled ICC stacks all 8 dimensions into a single matrix; per-dimension ICC is computed separately for each dimension. Neither panel shows a consistent advantage on per-dimension ICC; the flagship panel's primary benefit is substantially higher parse success rate.

#### Additional Data File

The real-case weight-sensitivity results are provided separately as `sensitivity-weights-real.json`. If the submission portal does not accept JSON supplementary files, this file should be converted to CSV or uploaded with a plain-text wrapper.
